# HCM-PhenotypeNet: Deep Multimodal Phenotyping of Hypertrophic Cardiomyopathy from Echocardiographic Video and Clinical Data

**DOI:** 10.64898/2026.09.13.26362954

**Authors:** Abdulsalam Almadani, Atifa Sarwar, Emmanuel Agu, Monica Ahluwalia, Jacques Kpodonu

## Abstract

**Background:** Hypertrophic cardiomyopathy (HCM) is a heterogeneous condition with variable morphology and symptoms, leading to adverse outcomes such as atrial fibrillation, heart failure and sudden cardiac death. Data-driven phenotyping may help refine risk stratification and guide management.

**Objectives:** To propose an automated machine learning framework (*HCM-PhenotypeNet*) that integrates echocardiographic video and clinical data to identify clinically meaningful HCM phenotypes.

**Methods:** We developed a multimodal pipeline using a deep neural network to extract features from 1,553 echocardiography video clips, combined with *>*100 clinical variables from 156 HCM patients. Patient-level vectors were reduced using UMAP and clustered using Deep Embedded Clustering (DEC). Cluster quality was assessed using internal validation metrics, and clusters were characterized through statistical comparisons and association rule mining.

**Results:** The cohort included 156 HCM patients (mean age 53.6 *±* 13.9 years, 40.4% female) with preserved systolic function and diverse racial/ethnic backgrounds. The optimal pipeline (DEC with 4 clusters) yielded phenogroups with strong separation (silhouette *≈*0.89): (A) Advanced Obstructive/Metabolic phenotype; (B) Early-onset, Genotype Positive phenotype; (C) Late-onset, mild non-obstructive phenotype; and (D) Hypertrophic Heart Failure-predominant phenotype. Whilst there was no mortality difference, there was a trend towards increased heart failure and atrial fibrillation burden in the Advanced Obstructive/Metabolic and Heart Failure-predominant subgroups.

**Conclusions:** *HCM-PhenotypeNet* enabled automated identification of four clinically relevant HCM subgroups. These phenogroups highlight the heterogeneity of HCM and may support personalized risk stratification and management but require validation in larger cohorts.

## 1. Introduction

Hypertrophic cardiomyopathy (HCM) is a genetic cardiac disease marked by unexplained myocardial thickening, often leading to heart failure (HF), arrhythmias, and less commonly sudden cardiac death (SCD). It is the most common inherited heart condition, with an estimated prevalence of about 1 in 500 in young adults based on echocardiographic screenings, though clinically diagnosed HCM is rarer (approximately 1 in 3,000) [1, 2]. The true prevalence is likely higher when including asymptomatic or undiagnosed cases.

Current HCM diagnostic criteria predominantly rely on a maximal end-diastolic wall thickness *≥*15 mm (by echocardiography or CMR) in the absence of other causes of hypertrophy [1, 3, 4, 5]. However, due to individual variation in gender, body size, and comorbidities, such strict cutoffs can result in overdiagnosis or underdiagnosis [1, 6]. Beyond wall thickness alone, other structural features (e.g., asymmetric septal hypertrophy, elongated mitral valve leaflets) also contribute to phenotypic expression. Recent guidelines emphasize detailed phenotyping to improve diagnosis and management of HCM [1]. *Phenotyping* involves classifying patients by meaningful clinical and imaging characteristics, such as patterns of left ventricular hypertrophy (LVH), electrical abnormalities, symptoms, or genetics rather than treating HCM as a single entity. Careful phenotyping enhances understanding of HCM’s diverse presentations and facilitates risk stratification, for example by identifying those at higher risk for adverse events like HF or SCD. It also lays the groundwork for personalized therapy: for instance, recognizing a phenotype with provocable left ventricular outflow tract (LVOT) obstruction would guide the use of cardiac myosin inhibitors and other standard therapies [1].

Prior work has explored unsupervised phenotyping in cardiovascular disease and HCM using predefined echocardiographic or latent-class approaches [7, 8, 9]. Our approach differs by leveraging deep learning on raw multi-view echocardiography videos integrated with structured clinical data, to perform automated HCM phenotyping. A side-by-side comparison of HCM phenotyping approaches is provided in Supplemental Table 1.

In this study, we propose *HCM-PhenotypeNet*, a novel multimodal framework that automatically discovers HCM patient subgroups by combining echocardiographic video analysis with structured clinical data. The core novelty of this work is threefold. First, phenotypic discovery is driven entirely by *raw pixel-level spatiotemporal sequences* inferred from multi-view echocardiographic videos—not by preprocessed traces, manually measured indices, or cardiologist-derived summary statistics, which are the input modalities utilized by all prior HCM unsupervised phenotyping studies [7, 8]. Second, the framework analyzes 1,553 video clips spanning four standard views (A2C, A4C, PLAX, PSAX), capturing complementary spatial perspectives of HCM morphology that no single-view analysis can provide. Third, raw video embeddings are fused with *>*100 structured clinical variables—including biomarkers, genetic testing results, and longitudinal outcomes—yielding a unified patient-level representation that enables holistic phenotyping that, to the best of our knowledge, has not been systematically demonstrated in HCM.

This framework also incorporates clinically annotated measures of cardiac function (e.g., mitral inflow velocity, systolic anterior motion, temporal LV wall motion patterns, as calculated by cardiologists), enabling a more holistic and automated characterization of HCM phenotypes than previous methods. Furthermore, by applying this framework to an ethnically diverse cohort (spanning 13 self-identified racial/ethnic categories), we address generalizability concerns since HCM expression can vary across populations [10]. We detail the development of *HCM-PhenotypeNet* and demonstrate how it identifies clinically meaningful HCM phenogroups that could improve risk stratification and management.

## 2. Methods

### 2.1. Study Population and Data Acquisition

We analyzed data from **HCM-Net**, a retrospective HCM cohort assembled in collaboration with the Boston Medical Center HCM program (IRB-approved, protocol H-44101). The dataset consists of 1,553 echocardiogram video clips (collected 2016–2021) from 167 individuals. Standard echocardiographic views were represented, including Apical 2-Chamber (A2C), Apical 4-Chamber (A4C), Parasternal Long Axis (PLAX), and Parasternal Short Axis (PSAX). Board-certified cardiologists confirmed HCM diagnoses based on clinical history, imaging findings, and genetic testing.

Diagnoses were made in accordance with the 2024 AHA/ACC HCM guidelines, which require unexplained LV hypertrophy (*≥*15 mm) in the absence of other loading conditions or systemic/metabolic diseases capable of producing the degree of hypertrophy observed (e.g., hypertensive cardiomyopathy, storage diseases, athlete’s heart, cardiac amyloidosis). Exclusion of phenocopies was based on clinical evaluation, echocardiographic morphology, and available genetic and metabolic workup as documented in the medical record.

Prior to clustering analyses, we excluded 11 patients who had incomplete demographics data or whose echocardiogram videos were misclassified as normal by the HCM-Dynamic-Echo model [11], which were unrelated to phenogroup assignment, yielding a final phenotyping cohort of 156 HCM patients. Over 100 structured clinical variables were curated per patient, including:

- **Demographics:** age at diagnosis, sex, BMI, ethnicity.
- **Cardiac measurements:** ejection fraction (EF), interventricular septal thickness (IVS), maximum wall thickness (MWT, CMR), chamber dimensions.
- **Additional data:** cardiac MRI findings (e.g., late gadolinium enhancement) and ECG findings (e.g., arrhythmias).
- **Clinical outcomes** included mortality (HCM-related death), NYHA class III–IV heart failure, left ventricular systolic dysfunction (LVEF *≤*35% and *≤*50%), and major cardiovascular events (AF, sustained ventricular tachycardia (VT), resuscitated arrest, cardioversion/appropriate ICD shocks). Additional outcomes were stroke/TIA, unplanned cardiac-related hospitalization, septal reduction therapies (septal myectomy and alcohol septal ablation), and ECMO, heart transplantation and/or LVAD. Follow-up was defined as the interval from the year of primary diagnosis (anchored to January 1 to standardize interval calculations across incomplete date records) to the most recent documented cardiology clinic visit, hospitalization, or death.

A comprehensive list of variables is provided in Supplemental Table 2. Representative frames from HCM and control subjects across four standard echocardiographic views are provided in Supplemental Table 3.

### 2.2. Phenotyping Pipeline (HCM-PhenotypeNet)

We developed the *HCM-PhenotypeNet* pipeline to identify phenotypic subgroups in hypertrophic cardiomyopathy by integrating echocardiographic video features and clinical data. An overview of the five-step pipeline is shown in Central Illustration

#### (1) Echocardiogram Video Feature Extraction

We utilized *HCM-Dynamic-Echo* [11], a deep learning model, to automatically extract a feature vector from each echocardiographic video. This model is based on the SlowFast video action recognition ar-chitecture [12], combining a slow branch that captures overall cardiac motion patterns across the cardiac cycle (e.g., progressive septal thickening, global LV systole and diastole) with a fast branch that detects very rapid events within a heartbeat (e.g., valve opening and closure, systolic anterior motion, or brief wall motion abnormalities). By integrating both global motion (semantically rich but temporally coarse) and rapid events (fine-grained temporal features), the model learns complementary representations of cardiac function. The model was pre-trained on the large EchoNet-Dynamic dataset (over 10,000 A4C echocardiogram videos from Stanford Medical School) [13] and fine-tuned on our HCM-Net videos, enabling robust feature learning specific to HCM.

To facilitate clinical inspection of phenogroup-level imaging patterns, representative A4C frames for each phenogroup are provided in Table 2, enabling uniform side-by-side visual comparison across all four clusters. It is important to note that phenogroup discovery is driven by spatiotemporal features learned jointly from all available multiview videos per patient, not from any single frame or view in isolation; thus, no individual still image fully captures the dynamic basis of cluster assignment.

#### (2) Feature Preprocessing and Aggregation

Each patient’s multiple video features were filtered for quality and then aggregated to form a single patient-level representation. We used *Connected Aggregation*, concatenating all echo video feature vectors for a patient:

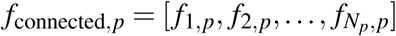

where *f_i,p_* is the feature from video *i* of patient *p*. This approach preserves within-patient heterogeneity and reduces dilution of infrequent yet clinically informative patterns. After aggregation, feature vectors were normalized to [0,1] using MinMax scaling. We then applied Uniform Manifold Approximation and Projection (UMAP) to reduce the 42-dimensional vectors to 2D embeddings, preserving local and global structure.

#### (3) Video Pattern Clustering

We applied unsupervised clustering to the UMAP embeddings to discover latent phenotypic groups. We selected Deep Embedded Clustering (DEC) as the primary unsupervised algorithm. DEC integrates an autoencoder-based latent representation with iterative clustering, refining both representation and assignments jointly. To determine the optimal number of clusters (*k*), we evaluated *k* values ranging from 2 to 10 using three internal validation metrics: the silhouette score, Calinski–Harabasz (CH) index, and Davies–Bouldin (DB) index. The final *k* was selected based on concordant improvement across these indices. Alternative clustering methods and definitions of validation metrics are provided in a Supplemental section on Clustering Algorithm Details.

#### (4) Association Analysis

Association analysis is a data science method used to discover co-occurrence or relationships between variables in large datasets. which we applied to discover which clinical variables were frequently associated with each HCM phenotype. For each variable, we performed ANOVA/Kruskal–Wallis or chi-square tests to assess variation across clusters (*p ≤* 0.05). In addition, frequent pattern mining (FP-Growth) [14] was applied within each cluster to identify high-confidence (*≥* 0.5) co-occurrences of clinical features. Continuous variables were binarized at prespecified clinical thresholds, and missing values were treated as not meeting the criterion (i.e., “absent” unless documented). Rules were retained when lift *≥*1.0 and interpreted as exploratory/hypothesis-generating. Full implementation details are provided in the Supplemental Association Rule Analysis.

#### (5) Phenogroup Definition

Phenogroups were labeled based on distinguishing features identified through statistical and association analyses. Each patient was assigned to one phenogroup for subsequent characterization.

## 3. Results

### 3.1. Baseline Characteristics

A total of 156 patients met inclusion criteria with a mean age at time of diagnosis of 53.6 *±* 13.9 years; 93 (59.6%) were male, mean BMI 30.2 *±* 6.4 kg/m^2^, 109 patients (70%) had hypertension based on past medical history, 43 (27.6%) had a history of diabetes mellitus (a key metabolic comorbidity distributed across phenogroups; see Table 1), 35 (22.4%) patients had a history of ICD placement. The mean left ventricular ejection fraction (LVEF) was 66.7 *±* 9.1% (n=155). The mean maximum wall thickness by CMR was 18.2 *±* 4.4 mm (n=94), and the mean left atrial size was 39.3 *±* 6.8 mm (n=142). Left ventricular outflow tract (LVOT) obstruction was present in 26 patients (16.7%) at baseline echo. The mean 5-year AHA/ACC sudden cardiac death risk score was 2.6 *±* 1.9 (n=155). Over a mean follow-up time of 6.4 *±* 6.4 years (median 4.6 years, IQR 2.0–8.0), atrial fibrillation occurred in 45 (29%) patients and sustained ventricular tachycardia or appropriate ICD therapy in 13 (8%) patients; overall, 79 (51%) had arrhythmias, 21 (13%) had TIA/stroke, 26 (17%) progressed to NYHA Class III–IV symptoms, 21 (13%) developed systolic dysfunction (LVEF *<*50%), 12 (7.7%) underwent septal reduction therapy, 2 (1%) underwent heart transplantation, 18 (11.5%) died (6 patients [3.8%] attributed to cardiac-related mortality).

**Table 1:**
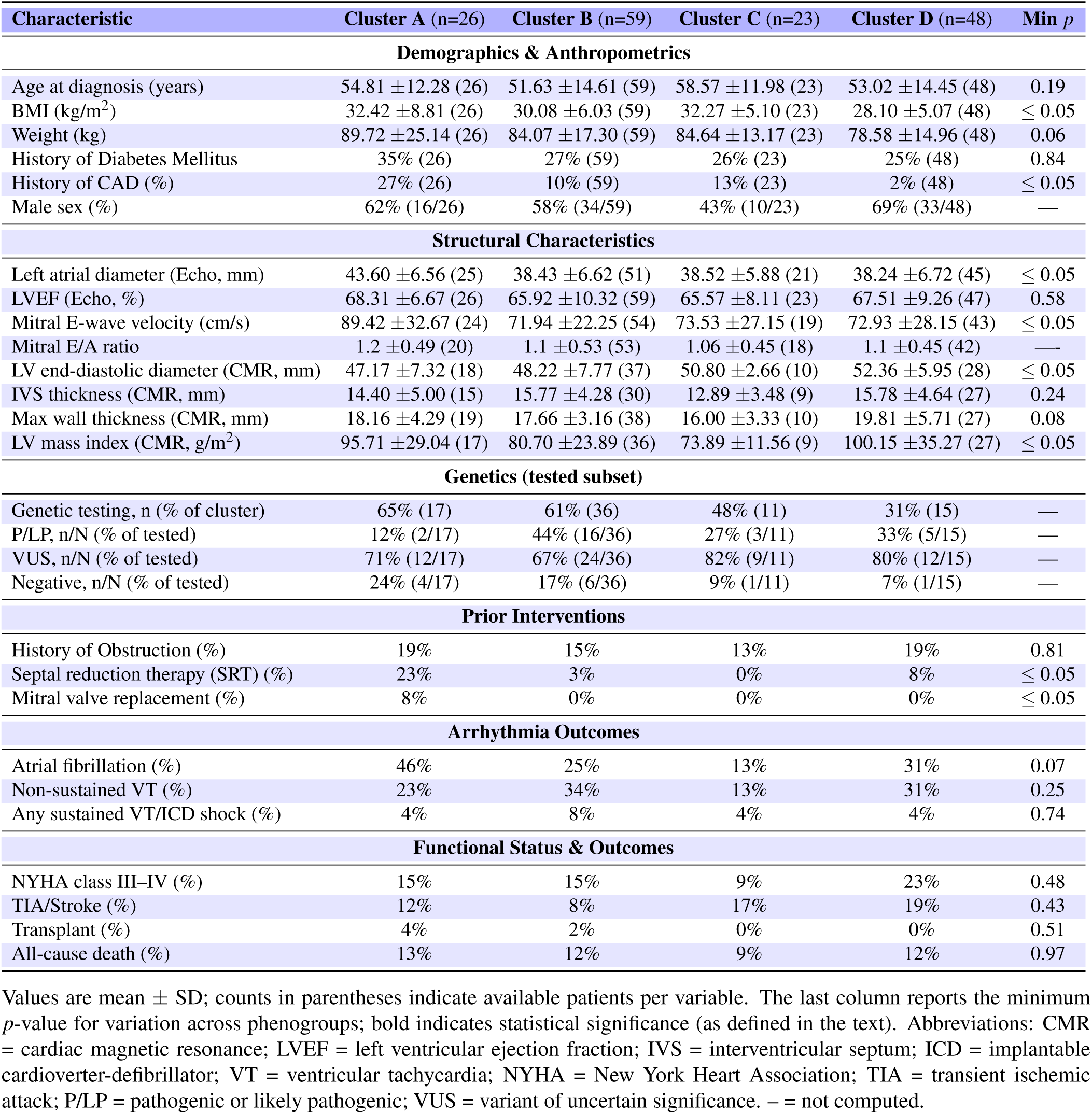
Clinical and structural features across HCM phenogroups.

| Characteristic | Cluster A (n=26) | Cluster B (n=59) | Cluster C (n=23) | Cluster D (n=48) | Min *p* |
| --- | --- | --- | --- | --- | --- |
| **Demographics & Anthropometrics** |  |  |  |  |  |
| Age at diagnosis (years) | 54.81 ±12.28 (26) | 51.63 ±14.61 (59) | 58.57 ±11.98 (23) | 53.02 ±14.45 (48) | 0.19 |
| BMI (kg/m<sup>2</sup>) | 32.42 ±8.81 (26) | 30.08 ±6.03 (59) | 32.27 ±5.10 (23) | 28.10 ±5.07 (48) | ≤ 0.05 |
| Weight (kg) | 89.72 ±25.14 (26) | 84.07 ±17.30 (59) | 84.64 ±13.17 (23) | 78.58 ±14.96 (48) | 0.06 |
| History of Diabetes Mellitus | 35% (26) | 27% (59) | 26% (23) | 25% (48) | 0.84 |
| History of CAD (%) | 27% (26) | 10% (59) | 13% (23) | 2% (48) | ≤ 0.05 |
| Male sex (%) | 62% (16/26) | 58% (34/59) | 43% (10/23) | 69% (33/48) | — |
| **Structural Characteristics** |  |  |  |  |  |
| Left atrial diameter (Echo, mm) | 43.60 ±6.56 (25) | 38.43 ±6.62 (51) | 38.52 ±5.88 (21) | 38.24 ±6.72 (45) | ≤ 0.05 |
| LVEF (Echo, %) | 68.31 ±6.67 (26) | 65.92 ±10.32 (59) | 65.57 ±8.11 (23) | 67.51 ±9.26 (47) | 0.58 |
| Mitral E-wave velocity (cm/s) | 89.42 ±32.67 (24) | 71.94 ±22.25 (54) | 73.53 ±27.15 (19) | 72.93 ±28.15 (43) | ≤ 0.05 |
| Mitral E/A ratio | 1.2 ±0.49 (20) | 1.1 ±0.53 (53) | 1.06 ±0.45 (18) | 1.1 ±0.45 (42) | — |
| LV end-diastolic diameter (CMR, mm) | 47.17 ±7.32 (18) | 48.22 ±7.77 (37) | 50.80 ±2.66 (10) | 52.36 ±5.95 (28) | ≤ 0.05 |
| IVS thickness (CMR, mm) | 14.40 ±5.00 (15) | 15.77 ±4.28 (30) | 12.89 ±3.48 (9) | 15.78 ±4.64 (27) | 0.24 |
| Max wall thickness (CMR, mm) | 18.16 ±4.29 (19) | 17.66 ±3.16 (38) | 16.00 ±3.33 (10) | 19.81 ±5.71 (27) | 0.08 |
| LV mass index (CMR, g/m<sup>2</sup>) | 95.71 ±29.04 (17) | 80.70 ±23.89 (36) | 73.89 ±11.56 (9) | 100.15 ±35.27 (27) | ≤ 0.05 |
| **Genetics (tested subset)** |  |  |  |  |  |
| Genetic testing, n (% of cluster) | 65% (17) | 61% (36) | 48% (11) | 31% (15) | — |
| P/LP, n/N (% of tested) | 12% (2/17) | 44% (16/36) | 27% (3/11) | 33% (5/15) | — |
| VUS, n/N (% of tested) | 71% (12/17) | 67% (24/36) | 82% (9/11) | 80% (12/15) | — |
| Negative, n/N (% of tested) | 24% (4/17) | 17% (6/36) | 9% (1/11) | 7% (1/15) | — |
| **Prior Interventions** |  |  |  |  |  |
| History of Obstruction (%) | 19% | 15% | 13% | 19% | 0.81 |
| Septal reduction therapy (SRT) (%) | 23% | 3% | 0% | 8% | ≤ 0.05 |
| Mitral valve replacement (%) | 8% | 0% | 0% | 0% | ≤ 0.05 |
| **Arrhythmia Outcomes** |  |  |  |  |  |
| Atrial fibrillation (%) | 46% | 25% | 13% | 31% | 0.07 |
| Non-sustained VT (%) | 23% | 34% | 13% | 31% | 0.25 |
| Any sustained VT/ICD shock (%) | 4% | 8% | 4% | 4% | 0.74 |
| **Functional Status & Outcomes** |  |  |  |  |  |
| NYHA class III–IV (%) | 15% | 15% | 9% | 23% | 0.48 |
| TIA/Stroke (%) | 12% | 8% | 17% | 19% | 0.43 |
| Transplant (%) | 4% | 2% | 0% | 0% | 0.51 |
| All-cause death (%) | 13% | 12% | 9% | 12% | 0.97 |

Values are mean ± SD; counts in parentheses indicate available patients per variable. The last column reports the minimum *p*-value for variation across phenogroups; bold indicates statistical significance (as defined in the text). Abbreviations: CMR = cardiac magnetic resonance; LVEF = left ventricular ejection fraction; IVS = interventricular septum; ICD = implantable cardioverter-defibrillator; VT = ventricular tachycardia; NYHA = New York Heart Association; TIA = transient ischemic attack; P/LP = pathogenic or likely pathogenic; VUS = variant of uncertain significance. — = not computed.

Figure 3.1 summarizes key demographic distributions.

### 3.2. Phenogroup Identification

Using our optimal configuration (Connected Aggregation + Normalization + UMAP + DEC at *k* = 4), *HCM-PhenotypeNet* identified four distinct patient clusters. These *phenogroups* were well separated in feature space (Figure 3.2) and corresponded to clinically recognizable profiles (described below). The silhouette score (0.89) indicates excellent cluster cohesion and separation.

For reference, Supplemental Figure 1 illustrates clustering performance metrics across varying configurations, confirming that the chosen pipeline outperformed alternatives in achieving high cluster separation.

All four phenogroups contained substantial numbers of patients (Cluster A: 26, B: 59, C: 23, D: 48), minimizing the chance that any cluster was a trivial outlier. Eight different clustering algorithms converged on these same four clusters, underscoring the robustness of the phenogroup structure.

As a further robustness check, we assessed the potential influence of prior Septal Reduction Therapy (SRT) on phenogroup assignments via a sensitivity analysis excluding the 12 patients with prior SRT. After excluding all 12 patients with prior septal reduction therapy (all from Phenogroup A), the four-cluster partition was fully preserved in the remaining cohort (n = 144), with high clustering quality metrics (silhouette score *≈*0.87; Calinski–Harabász index *≈*5,940; Davies–Bouldin index *≈*0.15). Additional details and results are reported in the Supplemental Material.

Multiple clustering configurations (*k* = 2–10) were systematically explored. The configuration with *k* = 4 consistently achieved the best balance across clustering quality metrics. Lower *k* values merged clinically distinct groups, whereas higher *k* fragmented stable phenogroups without improving interpretability. Thus, *k* = 4 was selected as the optimal balance between statistical robustness and clinical relevance. Detailed benchmarking of pipeline variations and clustering robustness is provided in Supplemental Results: Pipeline Performance and Alternative Configurations (Supplemental Figure 1).

### 3.3. Clinical Characteristics of Phenogroups

Each phenogroup exhibited a distinct constellation of structural and clinical features (Table 1). Features with significant associations (e.g., BMI, LA diameter, and mitral inflow velocity) were identified across phenogroups using standard statistical tests (ANOVA, Kruskal–Wallis, and Kolmogorov–Smirnov; see Table 1). The burden of arrhythmias, including atrial fibrillation, varied across phenogroups, suggesting differential electrical remodeling. The prevalence of obstruction was similar across phenogroups (A 19%, B 15%, C 13%, D 19%; p=0.81). A summary of key phenotypic traits and representative echocardiographic frames for each cluster is provided in Table 2. These phenogroup-level structural patterns are summarized visually in Figure 3.3. Expanded phenogroup narratives (Clusters A–D) are provided in the Supplemental Material (see radar plot, Supplemental Figure 2).

**Table 2:**
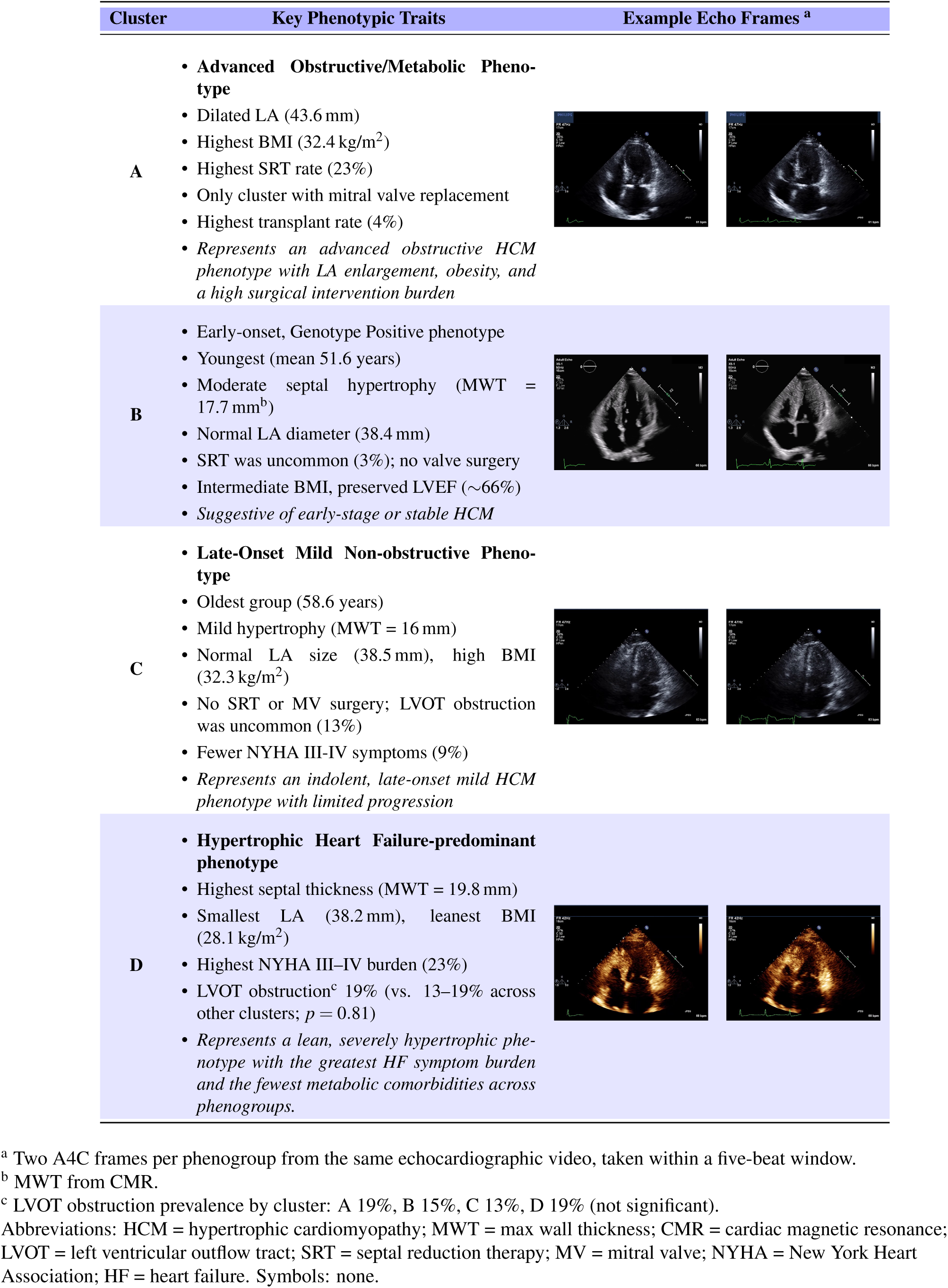
Phenotypic clusters in HCM with example echo frames.

| Cluster | Key Phenotypic Traits | Example Echo Frames <sup>a</sup> |
| --- | --- | --- |
| A | • **Advanced Obstructive/Metabolic Phenotype**<br>• Dilated LA (43.6 mm)<br>• Highest BMI (32.4 kg/m<sup>2</sup>)<br>• Highest SRT rate (23%)<br>• Only cluster with mitral valve replacement<br>• Highest transplant rate (4%)<br>• *Represents an advanced obstructive HCM phenotype with LA enlargement, obesity, and a high surgical intervention burden* |  |
| B | • Early-onset, Genotype Positive phenotype<br>• Youngest (mean 51.6 years)<br>• Moderate septal hypertrophy (MWT = 17.7 mm<sup>b</sup>)<br>• Normal LA diameter (38.4 mm)<br>• SRT was uncommon (3%); no valve surgery<br>• Intermediate BMI, preserved LVEF (~66%)<br>• *Suggestive of early-stage or stable HCM* |  |
| C | • **Late-Onset Mild Non-obstructive Phenotype**<br>• Oldest group (58.6 years)<br>• Mild hypertrophy (MWT = 16 mm)<br>• Normal LA size (38.5 mm), high BMI (32.3 kg/m<sup>2</sup>)<br>• No SRT or MV surgery; LVOT obstruction was uncommon (13%)<br>• Fewer NYHA III-IV symptoms (9%)<br>• *Represents an indolent, late-onset mild HCM phenotype with limited progression* |  |
| D | • **Hypertrophic Heart Failure-predominant phenotype**<br>• Highest septal thickness (MWT = 19.8 mm)<br>• Smallest LA (38.2 mm), leanest BMI (28.1 kg/m<sup>2</sup>)<br>• Highest NYHA III-IV burden (23%)<br>• LVOT obstruction<sup>c</sup> 19% (vs. 13–19% across other clusters; *p* = 0.81)<br>• *Represents a lean, severely hypertrophic phenotype with the greatest HF symptom burden and the fewest metabolic comorbidities across phenogroups.* |  |

<sup>a</sup> Two A4C frames per phenogroup from the same echocardiographic video, taken within a five-beat window.
<sup>b</sup> MWT from CMR.
<sup>c</sup> LVOT obstruction prevalence by cluster: A 19%, B 15%, C 13%, D 19% (not significant).
Abbreviations: HCM = hypertrophic cardiomyopathy; MWT = max wall thickness; CMR = cardiac magnetic resonance; LVOT = left ventricular outflow tract; SRT = septal reduction therapy; MV = mitral valve; NYHA = New York Heart Association; HF = heart failure. Symbols: none.

Across clusters, several structural and demographic metrics differed significantly (bold *p*-values in Table 1), while others (e.g., LVEF, sustained arrhythmias) did not. Notably, maximal wall thickness itself did not differ (*p* = 0.08), underscoring that the phenogroups reflect more than just hypertrophy severity. For example, Clusters A and C shared moderate MWT values but diverged markedly in BMI and intervention history.

No significant differences in major clinical outcomes were observed among clusters during follow-up median 4.6 years. Rates of all-cause death were similar (12.5% in each). Stroke/TIA incidence was also comparable across clusters (all *p >* 0.4). The lack of outcome divergence may reflect effective therapeutic interventions in higher-risk groups (e.g., ICD implantation, myectomy) or the need for longer follow-up and larger cohorts to detect differences. Importantly, the phenogroups still represent distinct phenotypic trajectories. For instance, patients in the Advanced Obstructive/Metabolic cluster (A) with larger left atria may face elevated long-term risks of atrial fibrillation or heart failure [8], while those in the Hypertrophic-HF cluster (D) with extreme hypertrophy may have increased arrhythmic burden, even if such differences were not apparent within our follow-up.

Thus, the four phenogroups identified by *HCM-PhenotypeNet* highlight meaningful heterogeneity in HCM. They differ significantly in cardiac structure and intervention history, despite similar survival in our cohort. This emphasizes that HCM is not monolithic—data-driven phenotyping can uncover subgroups that may benefit from tailored management. Differences in functional status and outcomes were also observed, with trends in NYHA class and heart failure hospitalizations providing additional clinical context (see Functional Status & Outcomes in Table 1).

## 4. Discussion

### 4.1. Advancing HCM Phenotyping with Multimodal Learning

Previous analyses have applied unsupervised learning to cardiovascular data to uncover disease subgroups. Loncaric *et al.* demonstrated that unsupervised clustering of whole-cycle echocardiographic tracings can identify functional phenotypes in heart disease, establishing the feasibility of ML-based phenotyping using dynamic echocardiographic data, though limited to hypertensive patients, excluding HCM [9]. In HCM, Loncaric *et al.* clustered manually derived echocardiographic and clinical features to define six phenogroups [7], while Marstrand *et al.* used latent class analysis of imaging and clinical variables to identify subtypes of obstructive HCM [8]. However, these approaches relied on pre-defined or externally derived echocardiographic measurements, utilized small cohorts, and did not leverage deep learning to analyze raw video data. Our *HCM-PhenotypeNet* extends these efforts by integrating raw multi-view echocardiographic videos with structured clinical data, enabling automated phenotyping (see Supplemental Table 1 for a detailed comparison).

Our results demonstrate that analyzing echo video features alongside clinical data can reveal distinct HCM phenotypes beyond those discovered by traditional classifications. Conventional HCM assessment relies on static measurements (like maximal wall thickness) or binary thresholds; in contrast, *HCM-PhenotypeNet* leverages dynamic imaging and multidimensional data to uncover subtle, latent patterns of disease expression. This approach represents a significant advance in precision cardiology, enabling identification of patient subgroups that might not be apparent from any single metric [9, 15]. By moving beyond one-size-fits-all criteria, our framework aligns with evolving guidelines advocating comprehensive early assessments of high-risk patients. It can potentially facilitate earlier recognition of high-risk profiles and better inform the timing of interventions.

### 4.2. Implications for Risk Stratification and Management

Identifying these four phenogroups has direct implications for patient care. Historically, HCM has been treated as a relatively homogeneous entity aside from differentiating obstructive vs non-obstructive forms. Our data-driven stratification reveals a spectrum of HCM phenotypes—from early-stage, relatively benign disease to advanced, HCM with evidence of more LA remodeling who require surgical intervention including septal reduction therapy and heart transplantation—similar to observations by Marstrand *et al.* that HCM encompasses diverse clinical trajectories [8]. Such stratification underscores the importance of individualized risk assessment and management.

These distinct phenotypic patterns (Table 2) highlight the heterogeneity of HCM and their potential implications for patient care. **Cluster A** reflected advanced obstructive/metabolic disease with atrial enlargement and obesity; **Cluster B** represented younger patients with moderate hypertrophy; **Cluster C** included older patients with milder disease ; and **Cluster D** corresponded to a Hypertrophic Heart Failure-predominant phenotype, defined by marked septal thickness (MWT 19.8 mm), lean body habitus (BMI 28.1 kg/m^2^), and the highest NYHA III–IV symptom burden (23%). Together, these groups suggest opportunities for phenotype-guided monitoring and management, complementing existing risk scores (e.g., 5-year SCD risk). The Hypertrophic HF label is descriptive rather than etiologic: it reflects the defining structural and symptomatic features of this group without presupposing a single genetic or morphologic mechanism.

Earlier studies by Loncaric *et al.* [7] recognized six phenotypic clusters among 138 participants (91 HCM patients and 47 relatives) by combining echocardiographic deformation and velocity traces with selected clinical features. Their approach demonstrated that unsupervised learning can uncover clinically relevant subtypes, including high-risk groups with severe hypertrophy and obstruction, intermediate phenotypes, and milder clusters enriched with relatives. However, these clusters were based on deformation and velocity profiles in a modest, single-country cohort, and ethnicity distribution was not reported. In contrast, *HCM-PhenotypeNet* uncovered four phenogroups in a larger cohort spanning multiple self-identified racial/ethnic categories, using deep learning to extract spatiotemporal features directly from *raw* echocardiographic videos and combining them with over 100 structured clinical variables. While Loncaric’s six clusters provided greater detail within a small dataset, our four phenogroups emphasize replicability, scalability, and generalizability, offering a complementary view of HCM heterogeneity. Similarly, Marstrand *et al.*[8] used latent class analysis in the SHaRe registry to identify clinically distinct subgroups of obstructive HCM, emphasizing that data-driven clustering can reveal outcome-significant subtypes. Together, these comparisons highlight that *HCM-PhenotypeNet* extends prior efforts by enabling fully automated, multimodal phenotyping across a diverse patient population.

### 4.3. Incremental Clinical Value Beyond Standard Echocardiographic Criteria

Standard HCM assessment relies principally on Maximal Wall Thickness (MWT *≥*15 mm) and the presence or absence of LVOT obstruction to guide risk stratification. *HCM-PhenotypeNet* identifies phenotypic structure that is not captured by these thresholds alone. Specifically, MWT did not significantly differ across the four phenogroups (*p* = 0.08, Table 1), yet the phenogroups diverged substantially in LA diameter, BMI, intervention burden, and arrhythmia trends—features with direct management implications. As a concrete example: Phenogroup D patients exhibit the highest MWT (19.8 mm) and the greatest NYHA III–IV symptom burden (23%), yet have the lowest BMI (28.1 kg/m^2^) and virtually absent CAD (2%)—a lean, hypertrophic, symptom-dominant profile that would be clinically indistinguishable from Phenogroup A by MWT threshold alone, yet carries a fundamentally different cardiometabolic risk profile and intervention trajectory. Similarly, Phenogroup C patients—the oldest group with the mildest hypertrophy (MWT 16 mm)—are at risk of being under-monitored under conventional criteria despite carrying atrial enlargement and metabolic comorbidities that may presage late-onset HF. By jointly leveraging dynamic video features and *>*100 clinical variables, *HCM-PhenotypeNet* surfaces these clinically actionable distinctions automatically, representing a meaningful increment over single-metric or binary threshold classification. These differences carry practical management implications. Phenogroup A (enlarged LA, high BMI, highest SRT rate) warrants heightened surveillance for AF and early discussion of septal reduction eligibility. Phenogroup B (youngest, moderate hypertrophy, no interventions) aligns with guideline-recommended annual monitoring and cardiac myosin inhibitor evaluation if obstruction develops. Phenogroup C (oldest, mildest hypertrophy) represents a profile at risk of under-monitoring despite LA enlargement; intensified rhythm monitoring and metabolic risk reduction may be warranted. Phenogroup D (extreme MWT, lean habitus, highest NYHA III–IV burden) suggests a symptomatic-dominant profile that may benefit from earlier optimization of heart failure pharmacotherapy and close arrhythmia surveillance despite low CAD burden. These suggestions are hypothesis-generating and require prospective validation before clinical adoption.

### 4.4. Genetic Testing and Phenotypic Correlations

Genetic testing was performed in 79 of 156 patients (50.6%), with unequal testing across phenogroups; genetic findings are therefore summarized descriptively. The genetic testing rate (*≈* 50%) aligns with current clinical practice. Detailed genotype distributions and thick-filament signal by phenogroup are provided in Supplemental Table 4 of the Supplemental materials.

Among tested patients, P/LP variants were more frequent in Phenogroups B and D (predominantly *MYBPC3*/*MYH7* thick-filament variants), whereas Phenogroups A and C showed VUS-dominant profiles with low sarcomeric signal (Supplemental Table 4). The Early-onset, Genotype Positive phenotype (Cluster B) was more likely to be genotype positive, which fits with established literature demonstrating that younger patients are more likely to be genotype positive. This pattern suggests that sarcomeric genetic burden may partially track phenogroup structure, though incomplete and unequal testing precludes formal association analysis.

### 4.5. Technical Insights and Robustness

Our analysis of many model variations revealed that the best results were consistently achieved by combining Connected Aggregation, UMAP, and DEC with *k* = 4. Notably, the same phenogroup structure was reproduced by many distinct clustering algorithms, underscoring the robustness of the underlying patient embeddings. We acknowledge the interpretive challenge inherent in deep learning–derived embeddings. To address this, phenogroup-representative echo frames (A4C view, applied uniformly across all four clusters) are provided in Table 2, enabling side-by-side visual comparison across clusters. The spatiotemporal feature space—learned jointly from slow (global cardiac motion) and fast (valve events, wall motion transients) branches of the SlowFast architecture—captures dynamic information not visible in static frames, which is precisely why multiview video analysis adds explanatory value beyond conventional echocardiographic measurements.

This robustness reflects that core phenotypic patterns were captured in the learned embeddings, independent of clustering algorithm. Unlike some prior unsupervised analyses, our careful design of feature extraction and dimensionality reduction yielded stable, clinically meaningful clusters regardless of the specific clustering method applied. This consistency indicates that our pipeline captures core phenotypic patterns that are not algorithm-dependent.

A clarification regarding model evaluation is warranted. *HCM-PhenotypeNet* performs *unsupervised* phenotype discovery rather than supervised outcome prediction; accordingly, there is no labeled ground truth against which a standard predictive baseline model could be benchmarked. Supervised models (e.g., logistic regression or gradient boosting on clinical variables) optimize for predicting a predefined outcome, which presupposes that meaningful subgroups and their clinical relevance are already known. In contrast, our framework uses internal clustering validity indices (silhouette, CH, and DB) as the appropriate evaluation framework for unsupervised partitioning, complemented by post-hoc statistical characterization of the discovered phenogroups. Standard echocardiographic measurements and clinical variables serve as interpretive anchors for phenogroup characterization—not as competing feature sets for a classification task. This distinction is fundamental to the phenotyping paradigm and aligns with established methodology in unsupervised cardiovascular phenotyping [7, 8].

### 4.6. Interpretable Patterns via Association Rules

Association rule mining was used as an interpretability layer to highlight within-phenogroup co-occurrence signatures (exploratory, hypothesis-generating). Complete rule definitions, metrics, and phenogroup-specific patterns are provided in the Supplemental Material (Association Rule Analysis; Supplemental Table 5).

### 4.7. Future Directions and Clinical Integration

Our study is a proof-of-concept demonstrating that ML-driven multimodal phenotyping can identify clinically meaningful HCM subgroups. Next steps include external validation on larger, multi-center cohorts and prospective studies to evaluate whether phenogroup membership adds prognostic value beyond traditional risk factors. A specific generalizability concern is the fine-tuning of the SlowFast feature extractor—pretrained on *>*10,000 general echocardiographic videos from a single institution (Stanford/EchoNet-Dynamic)—on our 156-patient, single-center HCM cohort. While transfer learning from a large echocardiographic dataset mitigates the risk of overfitting, the resulting embeddings may encode institution-specific acquisition characteristics (probe frequency, depth settings, frame rate) that limit portability to centers with different imaging protocols. External validation on independent HCM cohorts, ideally incorporating multiple acquisition systems and diverse racial/ethnic populations, is essential before clinical deployment. Full clinical integration considerations are provided in the Supplemental Material.

### 4.8. Study Limitations

This study has several limitations. It was retrospective and single-center, with modest sample sizes in some phenogroups (e.g., *n* = 26 in Cluster A), limiting power to detect outcome differences. Genetic test-ing was performed in only 50% of patients, which is a limitation. However, despite being a class I guideline recommendation, the real-world uptake of genetic testing in HCM probands is only approximately 57% based on a systematic review and meta-analysis of 48 studies [16]. This is comparable to what we have observed in our cohort. Because of this incomplete coverage across phenogroups, genetic yields are descriptive and may reflect ascertainment bias, and the possibility of phenocopies (e.g., storage diseases, cardiac amyloidosis) cannot be fully excluded in the untested subset, and automated cluster assignments were not systematically compared against confirmed genetic etiologies. The model was not tested on an independent external dataset, so generalizability remains to be established. The *HCM-Dynamic-Echo* feature extractor was fine-tuned on 156 patients at a single center; domain shift across institutions with different acquisition hardware and protocols may affect embedding transferability. Association rule analysis binarized continuous variables and emphasizes frequent patterns, potentially missing rare but clinically important associations. Finally, interventions (ICD implantation, myectomy) in higher-risk phenogroups may have attenuated detectable outcome differences during follow-up. A full methodological discussion of limitations is provided in the Supplemental Material.

## 5. Conclusions

We developed *HCM-PhenotypeNet*, a multimodal machine learning framework for automated hypertrophic cardiomyopathy phenotyping that integrates echocardiographic video analysis with structured clinical data. By combining deep learning-based video feature extraction with unsupervised clustering, *HCM-PhenotypeNet* moves beyond traditional diagnostic criteria (which rely on static, binary thresholds) to uncover distinct HCM subgroups that demonstrate clear differences in cardiac structure, electrophysiology, and clinical profiles [9, 8]. These data-driven phenogroups lay the groundwork for more refined risk stratification and personalized treatment plans—for example, one phenogroup represented advanced, obstructive HCM requiring invasive therapy, whereas another reflected a mild, latent onset HCM profile.

Our approach not only refines HCM classification but also offers a blueprint for integrating advanced computational analyses into routine practice. While challenges remain (e.g., in generalizability and workflow integration), the promising results reported here advocate for further exploration and validation of video-based phenotyping in HCM. Ultimately, this method represents a step toward precision medicine in cardiology, with the potential to transform HCM patient care through enhanced diagnostic accuracy and phenotype-guided management strategies.

## Supporting information

Main Text Figures

## Data Availability

All relevant clinical and analytical metrics used to support the study findings are included within the manuscript and its supplementary material. De-identified echocardiographic video data and electronic health record data are subject to institutional privacy policies and IRB restrictions at Boston Medical Center and are available from the corresponding author upon reasonable request for non-commercial research purposes.

## Acknowledgments

The authors thank the Boston Medical Center HCM Program and echocardiography laboratory staff Dr. Noyan Gocke and certified sonographers, Robert Cataldo and Jean Doricent, for their contributions to echocardiographic image acquisition. We also acknowledge the Worcester Polytechnic Institute Academic & Research Computing group for providing computational resources. Editorial assistance in reformatting the manuscript from a technical journal style to *JACC: Cardiovascular Imaging* was provided with the aid of ChatGPT (OpenAI, San Francisco, CA); all substantive content was authored and verified by the authors.

## Clinical Perspectives

### Competency in Medical Knowledge

HCM is a clinically heterogeneous disease. Using a combined deep learning echocardiography and clinical data approach, we identified four distinct HCM phenogroups with different structural and clinical characteristics. Recognizing these subgroups (e.g., an advanced obstructive and cardiometabolic phenotype with enlarged left atrium vs. Heart Failure-predominant phenotype) can help clinicians tailor monitoring and treatment to each patient’s specific risk profile.

### Translational Outlook

A multimodal phenotyping tool such as *HCM-PhenotypeNet* could be incorporated into echocardiographic analysis workflows to help stratify HCM patients at undergoing routine longitudinal care. Defining key HCM phenotypes including symptom burden may help provide more personalized management and may improve long-term outcomes. Further study with prospective validation in multi-center studies is needed.

## Abbreviations

AF: Atrial Fibrillation
BMI: Body Mass Index
EF: Ejection Fraction
HCM: Hypertrophic Cardiomyopathy
ICD: Implantable Cardioverter-Defibrillator
LA: Left Atrium
LV: Left Ventricle
LVEF: Left Ventricular Ejection Fraction
LVOT: Left Ventricular Outflow Tract
MWT: Maximal Wall Thickness

## Figure Legends

**Central Illustration.**
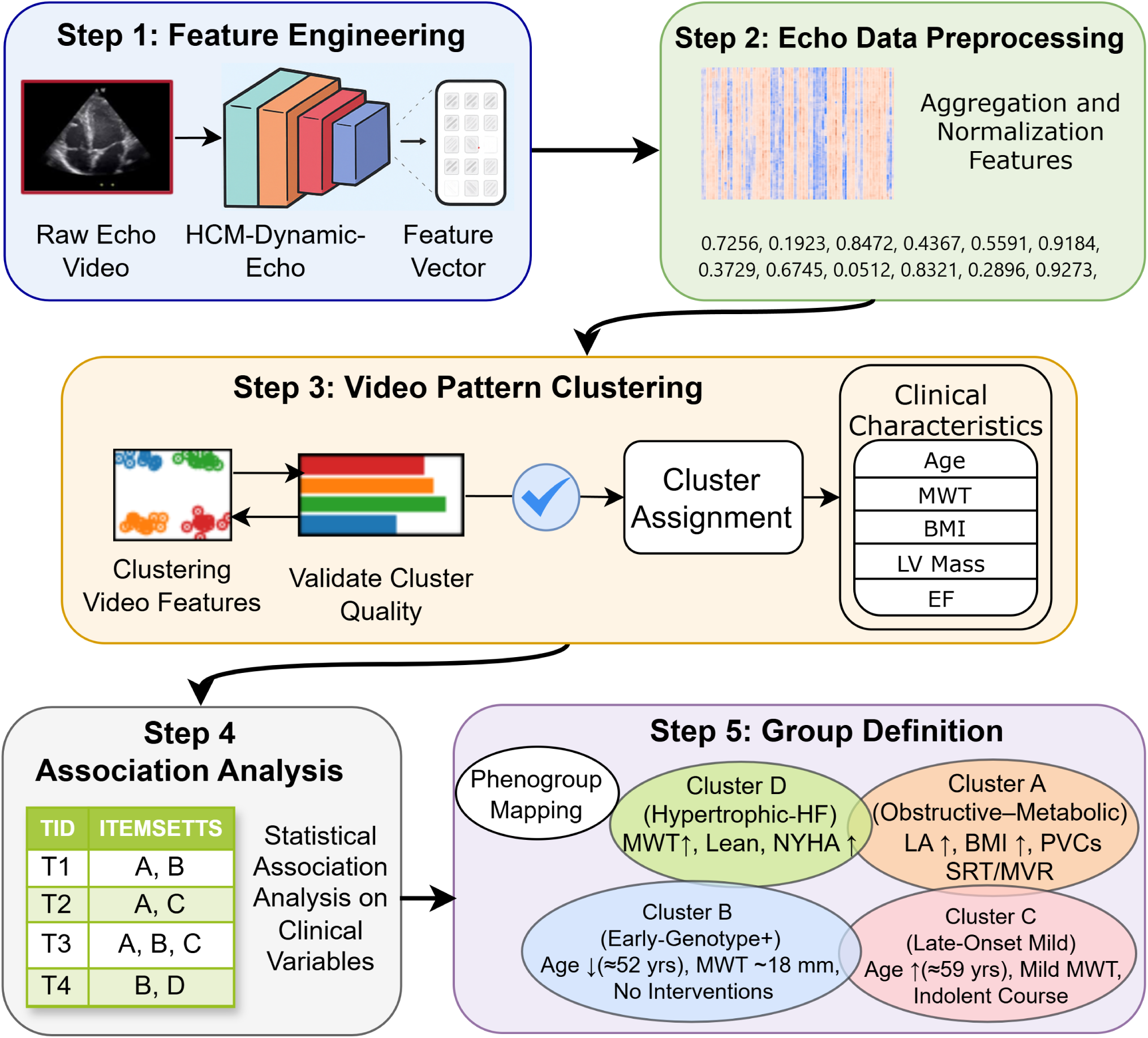
HCM-PhenotypeNet multimodal phenotyping workflow. Deep features are extracted from multi-view echocardiography videos and combined with clinical variables to form patient-level representations. These representations are embedded and clustered to assign phenogroups with internal validation. Phenogroups are then interpreted using statistical comparisons and association-rule mining to summarize defining clinical patterns. The five-step pipeline includes: (1) video feature engineering, (2) echo data preprocessing and aggregation, (3) video pattern clustering with quality validation, (4) association analysis of clinical variables, and (5) phenogroup definition and mapping. **Abbreviation:** HCM = hypertrophic cardiomyopathy.

**Figure 1.**
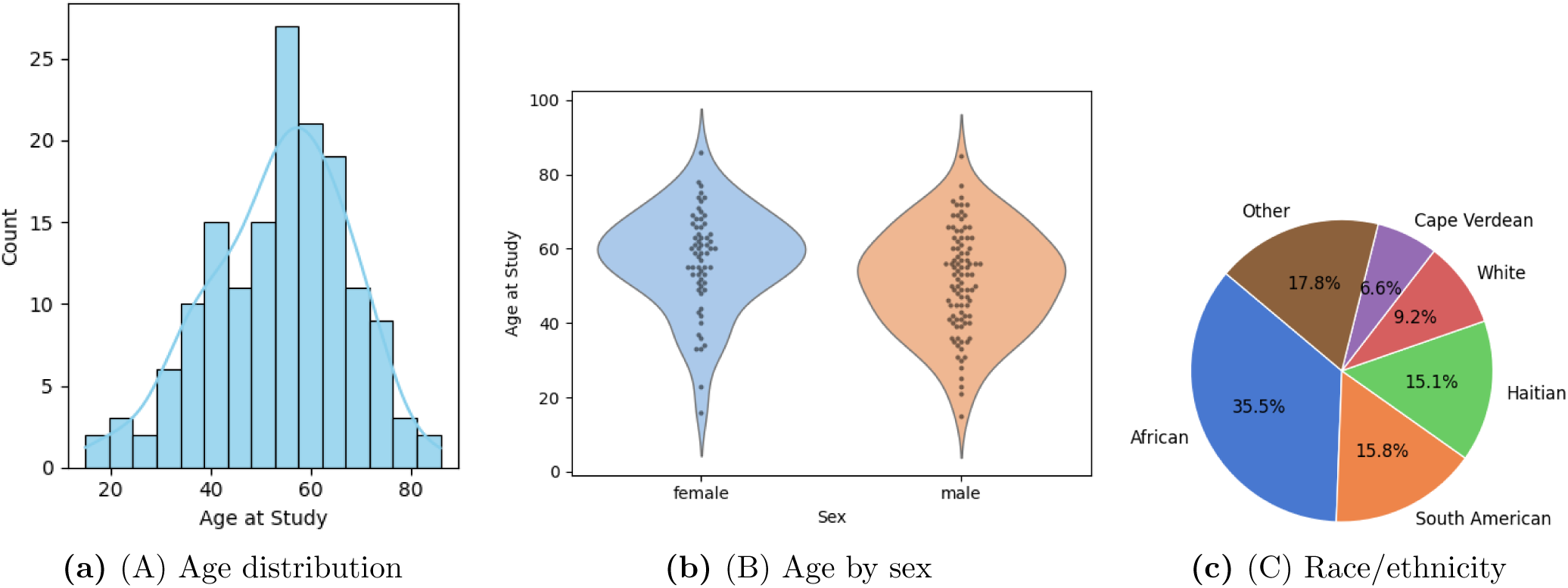
HCM-Net cohort demographics. Baseline demographic characteristics at the index echocardiogram are shown to contextualize downstream phenogroup comparisons. (A) Histogram of age at echocardiography with an overlaid kernel density estimate. (B) Violin plots show age distributions by sex. (C) Pie chart summarizes self-identified race/ethnicity (top five categories); remaining categories are grouped as “Other.”

**Figure 2.**
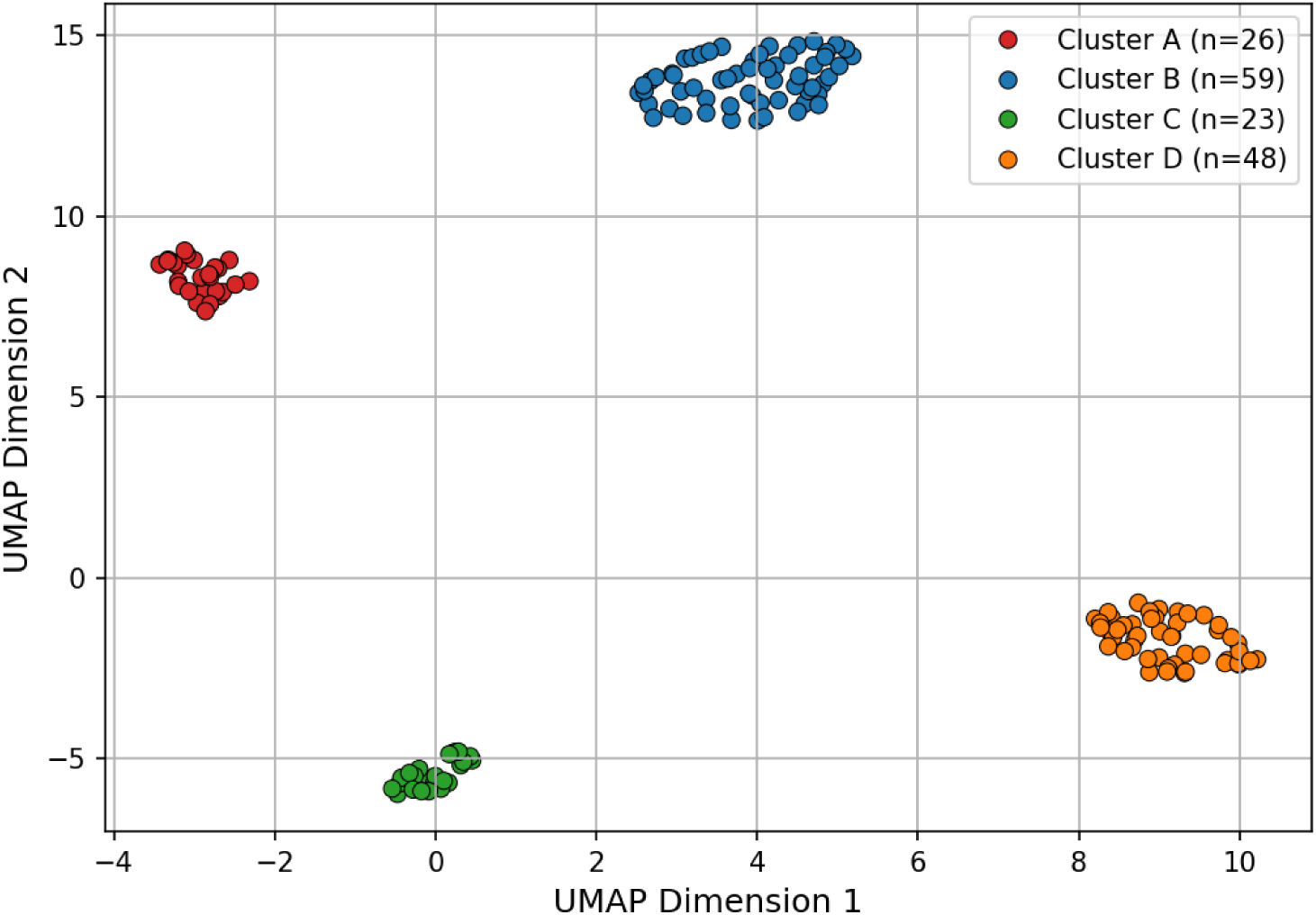
UMAP phenogroups in HCM. UMAP projection of patient-level embeddings for *k* = 4, where each point represents one HCM patient. Four clusters (A–D) define consensus phenogroups consistently recovered across the tested clustering algorithms. **Abbreviations:** UMAP = Uniform Manifold Approximation and Projection.

**Figure 3.**
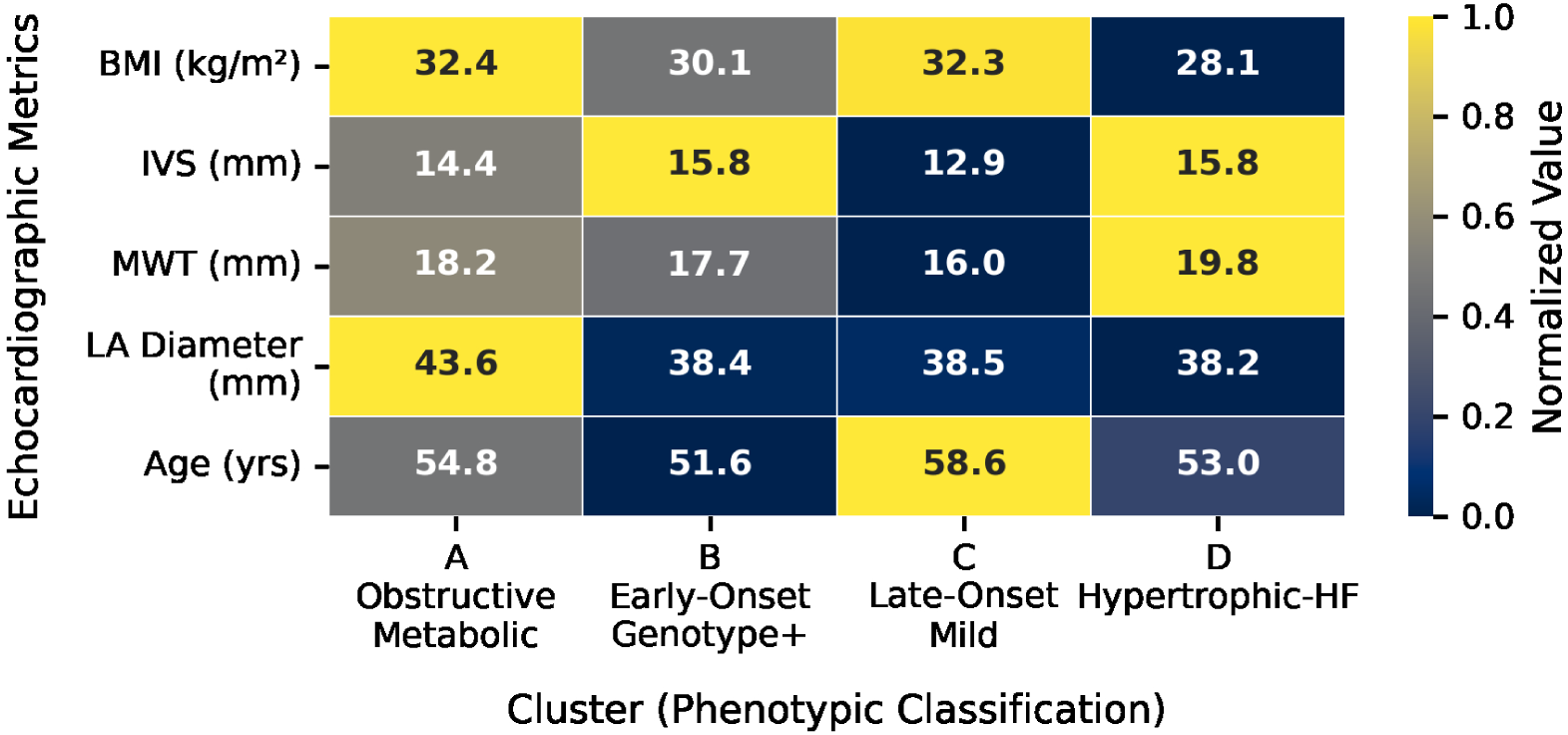
HCM-Phenogroup demographic heatmap. Normalized heatmap of echocardiographic and demographic metrics across the four phenogroups. Each row shows BMI, IVS thickness, MWT, LA diameter, and age, normalized by row. Lighter shading indicates higher mean values and darker shading lower mean values. Cluster A shows higher BMI and LA diameter; Cluster D shows higher MWT (CMR) but lower BMI. Phenogroup labels: Advanced Obstructive/Metabolic (A), Early-onset, Genotype+ (B), Late-Onset Mild Non-obstructive (C), and Hypertrophic-HF (D). A complementary radar plot is provided in Supplemental Figure 2. **Abbreviations:** BMI = body mass index; CMR = cardiac magnetic resonance; HCM = hypertrophic cardiomyopathy; IVS = interventricular septum; LA = left atrium; MWT = maximal wall thickness.

