## Supplementary material for "HCM-PhenotypeNet: Deep Multimodal Phenotyping of Hypertrophic Cardiomyopathy from Echocardiographic Video and Clinical Data": Main Text Figures

### HCM-PhenotypeNet: Figure Reference

All Main Manuscript Figures

Each figure is shown on its own page with full caption.

#### Contents

**1 Central Illustration — HCM-PhenotypeNet Workflow** **2**

**2 Figure 1 — HCM-Net Cohort Demographics** **3**

**3 Figure 2 — UMAP Phenogroups in HCM** **4**

**4 Figure 3 — HCM-Phenogroup Demographic Heatmap** **5**

**5 Quick-Reference Index** **6**

### 1 Central Illustration — HCM-PhenotypeNet Workflow

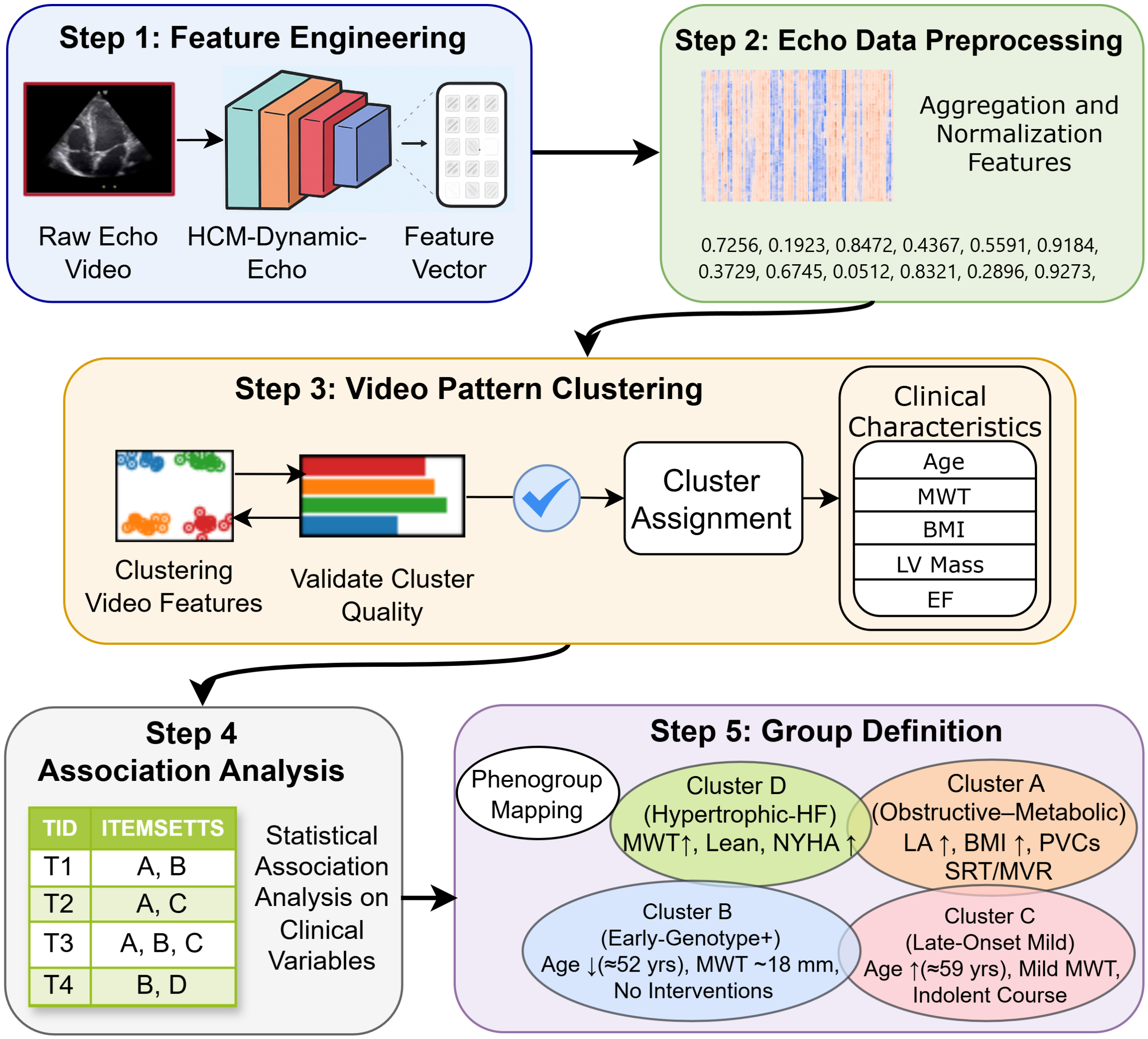


**Central Illustration. HCM-PhenotypeNet multimodal phenotyping workflow.** Deep features are extracted from multi-view echocardiography videos and combined with clinical variables to form patient-level representations. These representations are embedded and clustered to assign phenogroups with internal validation. Phenogroups are then interpreted using statistical comparisons and association-rule mining to summarize defining clinical patterns. The five-step pipeline includes: (1) video feature engineering, (2) echo data preprocessing and aggregation, (3) video pattern clustering with quality validation, (4) association analysis of clinical variables, and (5) phenogroup definition and mapping. **Abbreviation:** HCM = hypertrophic cardiomyopathy.

### 2 Figure 1 — HCM-Net Cohort Demographics

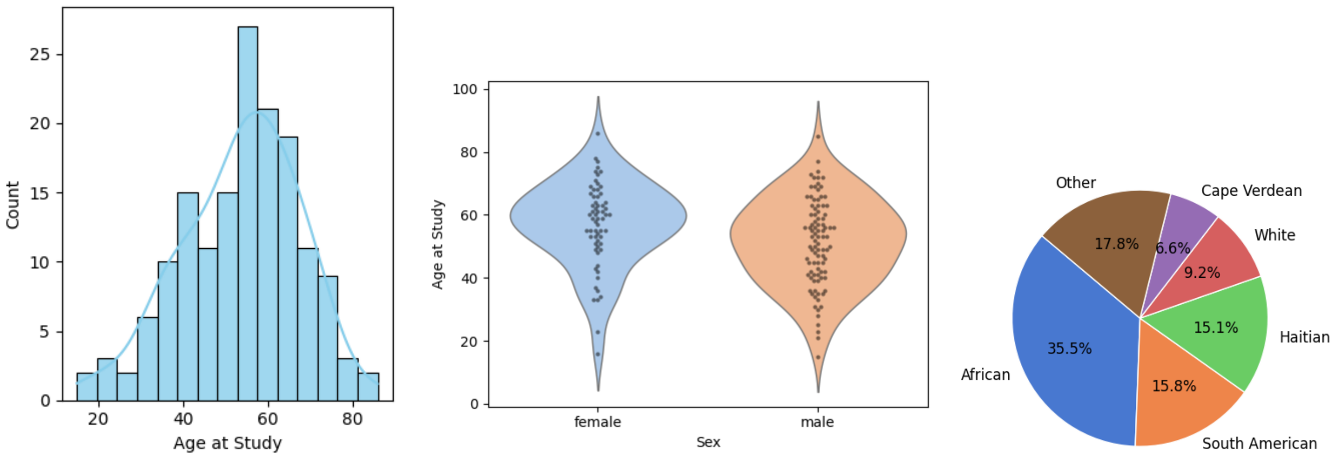


**(a)** (A) Age distribution **(b)** (B) Age by sex **(c)** (C) Race/ethnicity

**Figure 1: HCM-Net cohort demographics.** Baseline demographic characteristics at the index echocardiogram are shown to contextualize downstream phenogroup comparisons. (A) Histogram of age at echocardiography with an overlaid kernel density estimate. (B) Violin plots show age distributions by sex. (C) Pie chart summarizes self-identified race/ethnicity (top five categories); remaining categories are grouped as "Other."

### 3 Figure 2 — UMAP Phenogroups in HCM

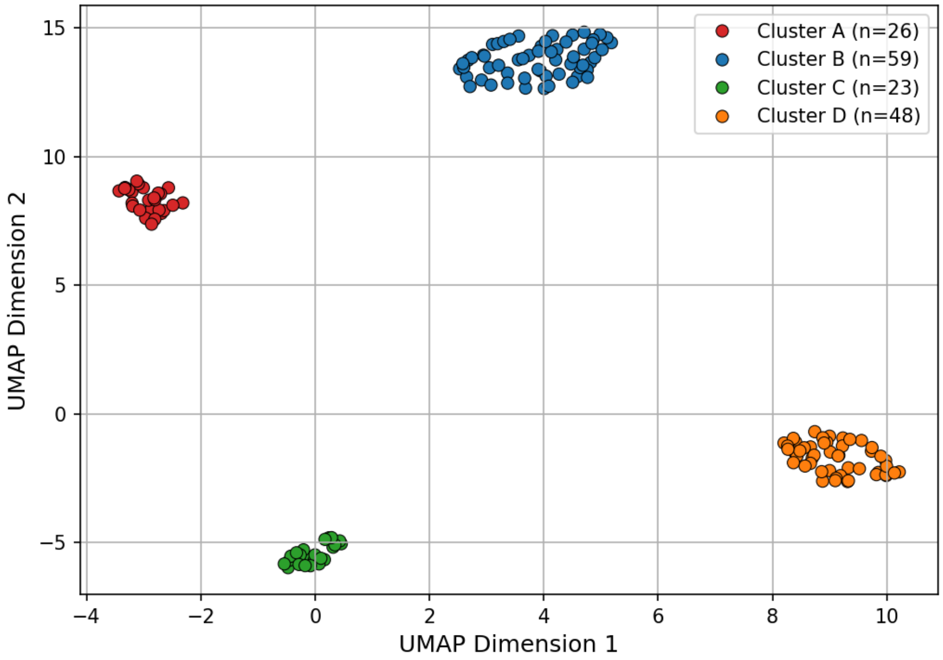


**Figure 2: UMAP phenogroups in HCM.** UMAP projection of patient-level embeddings for $k = 4$, where each point represents one HCM patient. Four clusters (A–D) define consensus phenogroups that were consistently recovered across the tested clustering algorithms. **Abbreviations:** UMAP = Uniform Manifold Approximation and Projection.

### 4 Figure 3 — HCM-Phenogroup Demographic Heatmap

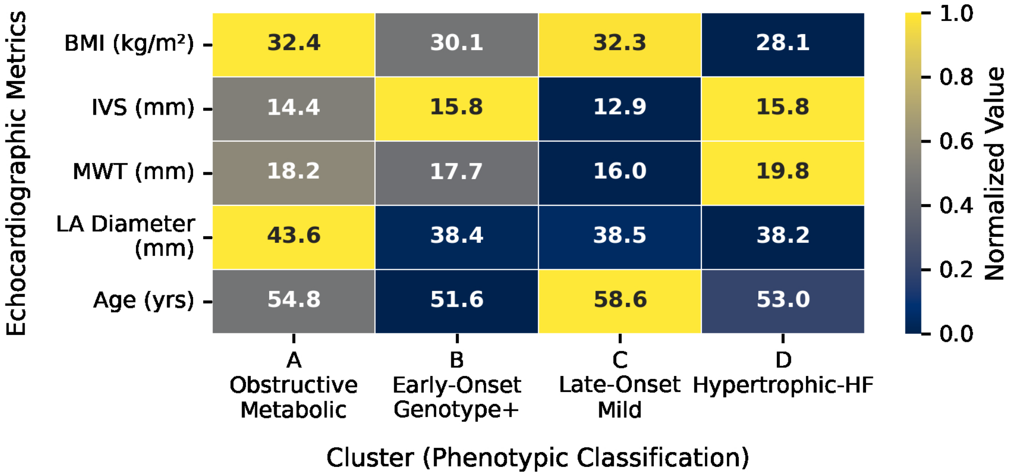


**Figure 3: HCM-Phenogroup demographic heatmap.** Normalized heatmap of echocardiographic and demographic metrics across the four phenogroups. Each row shows BMI, IVS thickness, MWT, LA diameter, and age, normalized by row. Lighter shading indicates higher mean values and darker shading lower mean values. Cluster A shows higher BMI and LA diameter, whereas Cluster D shows higher MWT (CMR) but lower BMI. Phenogroup labels reflect dominant traits: Advanced Obstructive/Metabolic (A), Early-onset, Genotype Positive (B), Late-Onset Mild Non-obstructive (C), and Hypertrophic Heart Failure-predominant (D). A complementary radar plot is provided in Supplemental Figure 2. **Abbreviations:** BMI = body mass index; CMR = cardiac magnetic resonance; HCM = hypertrophic cardiomyopathy; IVS = interventricular septum; LA = left atrium; MWT = maximal wall thickness.

### 5 Quick-Reference Index

| Figure | Source file | Caption summary |
|---|---|---|
| Central Illus. | `TIFF/Phase_2_steps.tiff` | 5-step HCM-PhenotypeNet pipeline diagram |
| Figure 1A | `demographic/TIFF/age_distribution_colab.tiff` | Age histogram with KDE |
| Figure 1B | `demographic/TIFF/age_by_sex_violin.tiff` | Age by sex violin plots |
| Figure 1C | `demographic/TIFF/race_ethnicity_pie_top5_plus_other.tiff` | Race/ethnicity pie chart |
| Figure 2 | `TIFF/features_list_connected_umap_dec_4.tiff` | UMAP $k$=4 cluster projection |
| Figure 3 | `TIFF/heatmap_clinical_from_latex.tiff` | Phenogroup demographic heatmap |
