## Supplementary Information for "HCM-PhenotypeNet: Deep Multimodal Phenotyping of Hypertrophic Cardiomyopathy from Echocardiographic Video and Clinical Data"

*Abdulsalam Almadani et al.*

---

#### Supplemental Introduction

*Supplemental Review of Related Studies*

**Loncaric et al**. [1] and **Marstrand et al.** [2] studies highlight the feasibility of unsupervised phenotyping in HCM but were constrained by reliance on predefined measurements and limited cohort sizes. Supplemental Table 1 presents a structured comparison of their methods and our HCM-PhenotypeNet framework.

#### Supplemental Methods

Supplemental Note S2. Additional Methods Details

*Study Population and Data Acquisition (Supplemental)*

To complement the main manuscript, we provide additional structured data and representative imaging frames from the HCM-Net cohort.

*Clustering Algorithm Details*

For completeness, we detail the clustering algorithms and metrics evaluated in our study. We compared DEC (used in the main analysis) to a range of clustering methods:

- **K-Means:** Partitions data into $k$ clusters by minimizing intra-cluster variance. It seeks to minimize $\sum_{i=1}^{k}\sum_{x\in S_i}\|x-\mu_i\|^2$, where $\mu_i$ is the centroid of cluster $S_i$. K-means is efficient for well-separated spherical clusters but can perform poorly if clusters have complex shapes or different sizes [3, 4].
- **Agglomerative Hierarchical Clustering:** Builds a hierarchy of clusters by iteratively merging the closest pairs. It can reveal nested subgroup structures, but is less practical for very large datasets due to its $O(n^2)$ complexity [5].

Supplemental Table 1: Comparison of Phenotyping Strategies in Related Studies vs. Our Work.

| Aspect | Loncaric et al. 2021 [1] | Marstrand et al. 2020 [2] | HCM-PhenotypeNet (Ours) |
|---|---|---|---|
| **Cohort Size and Composition** | 138 participants (91 HCM, 47 relatives), Spain, presumed limited ethnic diversity | ∼300 obstructive HCM patients from Danish cohort | 156 HCM patients with 1,553 echocardiogram videos, multi-ethnic U.S. population, single center |
| **Input Modality** | Whole cardiac-cycle echo deformation + clinical data | Echo parameters and clinical measurements | Raw multi-view echo videos + structured clinical data |
| **Feature Extraction** | Manually extracted echo traces + clinical variables | Manually measured imaging and clinical features | Deep learning (SlowFast architecture) for video feature extraction |
| **Clustering Method** | Unsupervised clustering in ML-derived latent space (6 phenogroups) | Latent class analysis | Deep Embedded Clustering (DEC) |
| **Dimensionality Reduction** | ML-derived low-dimensional embedding (autoencoder) | Not applicable (no separate embedding) | UMAP (nonlinear manifold embedding) |
| **Modality Integration** | Manual integration of echo traces and clinical variables | Manual integration of echo and clinical data | Multimodal: echo video features + >100 clinical variables combined |
| **Phenotyping Focus** | Functional and clinical subgroups in HCM patients and relatives | Clinical stratification of obstructive HCM | Spatiotemporal echo-based phenotyping of HCM |
| **Interpretability** | Clinical and genetic profiles of clusters described | Survival outcomes and clinical profiles of clusters | Statistical associations of phenogroups with clinical variables (e.g., interventions, risk factors) |

- **Spectral Clustering:** Constructs a similarity graph of the data and uses eigen-decomposition of its Laplacian to perform dimensionality reduction before clustering (often via k-means) [6]. It captures non-convex cluster shapes well, but performance depends on the graph construction parameters.
- **BIRCH (Balanced Iterative Reducing and Clustering using Hierarchies):** An incremental clustering that builds a tree of cluster feature summaries (CF-tree) to efficiently cluster large datasets [7]. It trades some accuracy for speed and memory efficiency and may struggle if cluster densities vary.

Supplemental Table 2: Clinical and Echocardiographic Variables Collected in HCM-Net Dataset.

| **Clinical Characteristics** | **Echocardiographic Variables** | **Clinical Outcomes** |
|---|---|---|
| Age at diagnosis (years) | Maximal LV wall thickness (mm) | Overall composite |
| Sex | LV mass indexed per body surface area (g/m²) | NYHA Class III-IV |
| Body mass index (kg/m²) | Left atrial (LA) diameter | HF Hospitalization |
| Sarcomere positive genotype | Left atrial volume (cc/m²) | Incident Atrial Fibrillation |
| Race | LVEF $< 50\%$ | Stroke |
| Social Determinants of Health | LVOT peak gradient $\geq$ 30 mmHg | Appropriate ICD therapy |
| New York Heart Association Class | Septal e' (cm/s) | Septal reduction therapy |
| Systolic Blood Pressure (mmHg) | Lateral e' (cm/s) | VAD/Transplant |
| History of syncope | Global longitudinal strain (%) | Sudden cardiac death |
| Family history of sudden cardiac death | Systolic anterior motion of mitral valve | HF composite outcome |
| Non-sustained ventricular arrhythmia | Significant mitral regurgitation (3+) | NYHA Class III-IV |
| Troponin T (ng/mL) | | HF Hospitalization |
| N-terminal pro-B-type natriuretic peptide (pg/mL) | | VAD/Transplant |
| Late gadolinium enhancement on cardiac MRI (%) | | |
| Apical aneurysm on cardiac MRI | | |
| ESC SCD risk score | | |
| Disopyramide use | | |
| Myosin inhibitor use | | |

- **MiniBatch K-Means:** A faster variant of k-means that uses small random batches to update centroids [8]. It scales well but can yield less stable clusters due to its stochastic nature.

- **DBSCAN (Density-Based Spatial Clustering of Applications with Noise):** Defines clusters as regions of high density separated by low-density areas [9]. It can find arbitrarily shaped clusters and identify outliers, but requires tuning a density threshold and may merge clusters if densities are similar.

- **OPTICS (Ordering Points to Identify Clustering Structure):** A density-based method related to DBSCAN that outputs an ordering (reachability plot) illustrating cluster structure [10, 11]. It handles varying density clusters better but is more complex to interpret.

- **Affinity Propagation:** Does not require a pre-set $k$. It finds "exemplar" points by iterative message passing between all pairs of points [12]. It can automatically determine cluster count, but its $O(n^2)$ complexity and sensitivity to parameters limit its practical use.

- **MeanShift:** A non-parametric clustering that shifts each point towards the local density peak using a kernel density estimate [13]. It can find arbitrarily shaped clusters and determine their number, but is computationally intensive for large datasets.

Supplemental Table 3: Representative Echocardiographic Frames From an HCM Patient and a Control Subject Across Four Standard Views (A2C, PLAX, PSAX, A4C)

| View | HCM | Control |
|---|---|---|
| A2C | 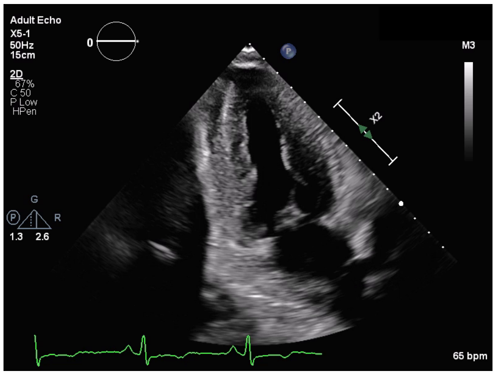 | 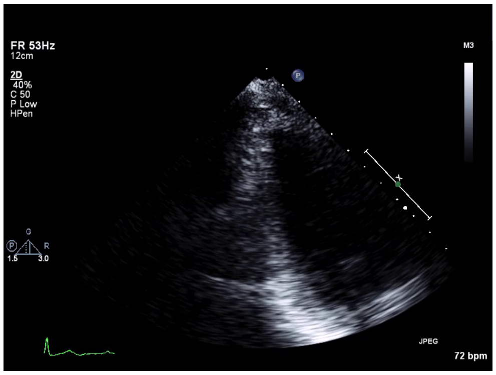 |
| PLAX | 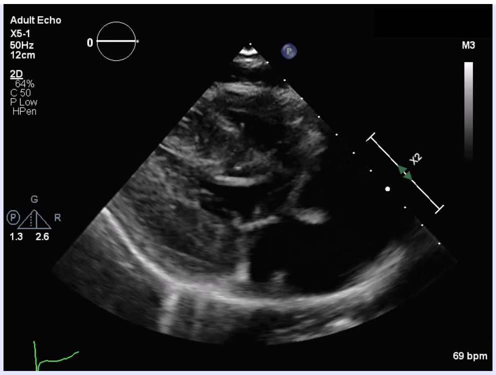 | 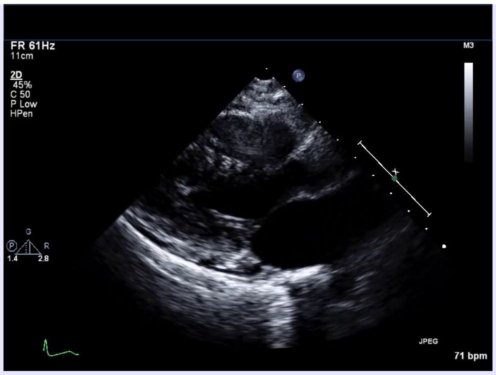 |
| PSAX | 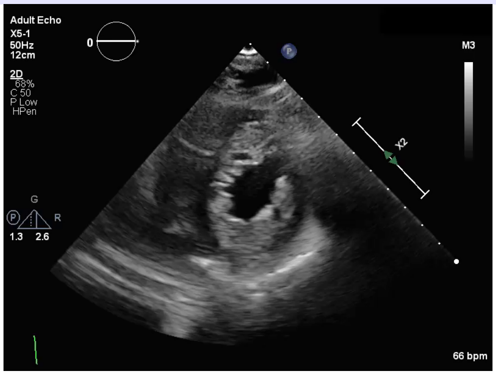 | 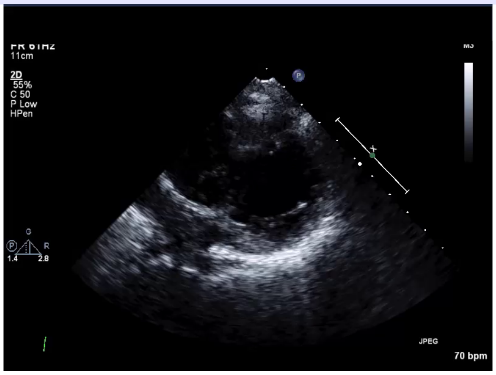 |
| A4C | 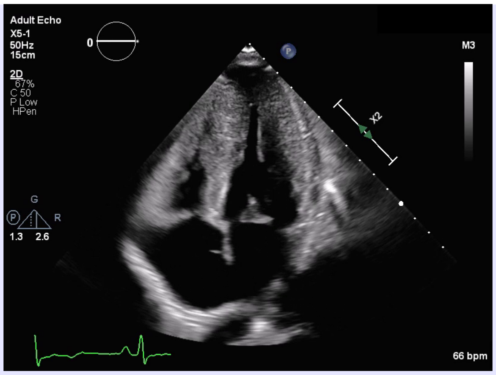 | 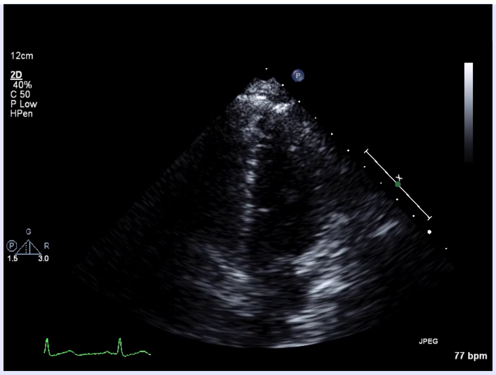 |

Across these methods, the same four clusters consistently emerged (excluding methods like OPTICS/DBSCAN when they did not require a fixed $k$). This underscores the intrinsic separability of our data.

*Clustering Quality Metrics*

We utilized three metrics to evaluate cluster quality:

1. **Silhouette Score:** For each point, compute $a =$ average distance to other points in its cluster, and $b =$ minimum average distance to points in any other cluster. The silhouette $S = (b-a)/\max(a,b)$. It ranges from $-1$ to 1; higher values indicate better-defined clusters. In our data, the mean silhouette $\approx 0.89$, indicating clusters are well separated with little overlap.

2. **Calinski–Harabasz (CH) Index:** Ratio of between-cluster to within-cluster variance: $\mathrm{CH} = \frac{\mathrm{trace}(B_k)/(k-1)}{\mathrm{trace}(W_k)/(N-k)}$, where $B_k$ and $W_k$ are the between- and within-cluster dispersion matrices, $N$ is total points and $k$ cluster count. Higher CH suggests compact, well-separated clusters. In our results, CH was $\approx 6241.9$, very high due to large between-group variance.

3. **Davies–Bouldin (DB) Index:** Measures average similarity between clusters, defined as $\frac{1}{k}\sum_{i=1}^{k}\max_{j\neq i}\frac{\sigma_i+\sigma_j}{d_{ij}}$, where $\sigma_i$ is cluster $i$'s average distance to its centroid and $d_{ij}$ distance between centroids of clusters $i$ and $j$. Lower DB indicates better separation (clusters far apart relative to their size). Our DB $\approx 0.1408$, close to 0, reflecting very well-separated clusters.

### 1. Supplemental Results

Expanded Phenogroup Clinical Narratives (Clusters A–D) (Main Results 3.3)

- **Cluster A** ("Advanced Obstructive/Metabolic" phenotype) had the largest mean left atrial (LA) diameter (43.6 mm vs. 38 mm in others, $p \leq 0.05$) and highest mean BMI ( 32.4 kg/m$^2$, $p \approx 0.01$). It also had the greatest burden of coronary artery disease (CAD) (27% vs. 2% in Cluster D, $p \leq 0.05$) and the highest prevalence of diabetes (35 % vs $\approx$ 25–27% in others). Importantly, 23% of Cluster A patients had undergone septal reduction therapy (SRT) (versus $\leq$ 8% in others, $p \approx 0.006$), and 8% had mitral valve replacement (0% in others). This cluster also had the highest transplant rate (4% vs. $\leq$2% in others). This phenogroup thus represents advanced obstructive HCM—characterized by left atrial enlargement, obesity, left ventricular outflow tract obstruction, and concomitant mitral valve disease requiring a high rate of surgical interventions.

- **Cluster B** Early-onset, Genotype Positive phenotype patients were the youngest on average (mean 51.6 years). They had moderate septal hypertrophy (MWT 17.7 mm) and relatively smaller LA diameter (38.4 mm). BMI was intermediate (mean 30 kg/m$^2$). Prior SRT was uncommon (3%); no valve surgery. This group may reflect earlier-stage HCM with moderate structural changes.

- **Cluster C** ("Late-Onset Mild Non-obstructive HCM" phenotype) was the oldest group (mean 58.6 years) and had the mildest hypertrophy (MWT 16 mm; IVS  13.9 mm). LA size (38.5 mm) and BMI (32.3 kg/m$^2$) were similar to others. Notably, none had undergone septal reduction or mitral surgery. They also had the fewest advanced HF symptoms (only 9% NYHA III–IV vs. 23% in Cluster D). This cluster likely represents an indolent, late-onset mild HCM phenotype characterized by smaller LA size and fewer advanced symptoms, occurring later in life with relatively mild structural remodeling.

- **Cluster D** ("Hypertrophic Heart Failure-predominant phenotype") included features across the HCM spectrum, with some obstructive cases ($\approx$19%), rather than a primarily obstructive cohort. It showed the highest septal thickness (MWT 19.8 mm) but leanest body habitus (mean BMI 28.1 kg/m$^2$). This cohort had the smallest LA diameter (38.2 mm) and included more symptomatic patients (23% NYHA III–IV, though $p = 0.48$ vs. others). About 19% of patients had LVOT obstruction, of whom

8% underwent SRT; the remainder were managed medically. This phenogroup represents a relatively hypertrophic HF-predominant phenotype (lowest BMI) with greater heart failure symptom burden (highest proportion of NYHA III–IV), modest obstruction, and fewer metabolic comorbidities (markedly lower CAD and slightly lower diabetes prevalence).

A complementary radar plot summarizing normalized phenogroup features is shown in Supplemental Figure 2.

*Genetic Testing and Phenotypic Correlations*

This section expands on the main manuscript Section 4.3.

Genetic testing was performed in 79 of 156 patients (50.6%), with phenogroup coverage ranging from ($\approx$ 31–65%) (Main Table 1; Supplemental Table 4). Given partial and unequal testing, genetic results are summarized descriptively. Thick-filament involvement refers to variants in *MYBPC3*/*MYH7*. Among those tested, pathogenic or likely pathogenic (P/LP) variants were more frequent in cluster B and D, whereas variant of uncertain significance (VUS) findings predominated in cluster A and C. Phenogroup A showed low P/LP yield and sparse thick-filament signal, B had the highest sarcomeric burden (commonly *MYBPC3*/*MYH7*) often alongside non-sarcomeric VUS, C showed modest P/LP and stacked sarcomeric VUS with age-modulated expression and D had intermediate P/LP with clear sarcomeric signal. Practically, a P/LP anchor (notably in B and D) supports cascade testing and genotype-informed counseling, whereas VUS-dominant profiles (A and C) favor phenotype-guided management and periodic variant reclassification.

*Pipeline Performance and Alternative Configurations*

We systematically explored various feature aggregation, normalization, and dimensionality reduction options. Supplemental Figure 1 summarizes clustering performance (silhouette, CH, DB indices) for different numbers of clusters ($k = 2$ to 10) across configurations. The combination of Connected Aggregation + MinMax scaling + UMAP + DEC at $k = 4$ achieved the best overall metrics (highest silhouette & CH; lowest DB). Alternative configurations (e.g., using PCA instead of UMAP, or mean pooling instead of concatenation) yielded either lower cluster separation or unstable cluster structures.

Notably, clustering outcomes were robust across algorithmic families when using our optimal pipeline's embedding. Eight of ten clustering algorithms (all except methods that define their own optimal $k$ like

Supplemental Table 4: Genetic Signal by Phenogroup (Tested Subset).

| Phenogroup | Sarc+ (n) | Sarc–VUS (n) | Sarc– (n) | Thick-filament signal[a] |
|---|---|---|---|---|
| A | 2 | 4 | 10 | 1 P/LP patients; 2 VUS patients; 2 VUS calls |
| B | 13 | 7 | 17 | 13 P/LP patients; 4 VUS patients; 4 VUS calls |
| C | 3 | 4 | 4 | 3 P/LP patients; 3 VUS patients; 5 VUS calls |
| D | 4 | 4 | 8 | 4 P/LP patients; 3 VUS patients; 3 VUS calls |

[a] Thick-filament signal summarizes MYBPC3/MYH7 involvement as: P/LP patients; VUS patients; total VUS calls. Counts shown for sarcomeric P/LP (Sarc+), sarcomeric VUS (Sarc–VUS), and no sarcomeric variant (Sarc–). Representative P/LP variants are listed. Phenogroups: A = Advanced Obstructive/Metabolic; B = Intermediate Non-obstructive; C = Late-Onset Mild Non-obstructive; D = Hypertrophic HF-predominant phenotype. **Representative P/LP examples (non-exhaustive):** A: TNNT2 p.Trp287*, MYBPC3 c.111del (p.Val38*). B: MYBPC3 p.Arg502Trp; splice donor c.821+1G>A; MYH7 p.Arg1820Gln. C: MYBPC3 p.Glu258Lys; MYL3 p.Glu143Lys. D: MYBPC3 p.Trp322*; p.Asp770Asn.
**Abbreviations:** HCM = hypertrophic cardiomyopathy; MYBPC3 = myosin binding protein cardiac type 3; MYH7 = beta-myosin heavy chain 7; P/LP = pathogenic or likely pathogenic; Sarc+ = sarcomeric P/LP; Sarc– = no sarcomeric variant detected; Sarc–VUS = sarcomeric variant of uncertain significance; TNNT2 = troponin T2; VUS = variants of uncertain significance.

Affinity Propagation and OPTICS) converged on identical 4-cluster assignments. This agreement highlights the stability of the patient feature representation we derived.

**Supplemental Figure 1 Clustering performance in HCM**

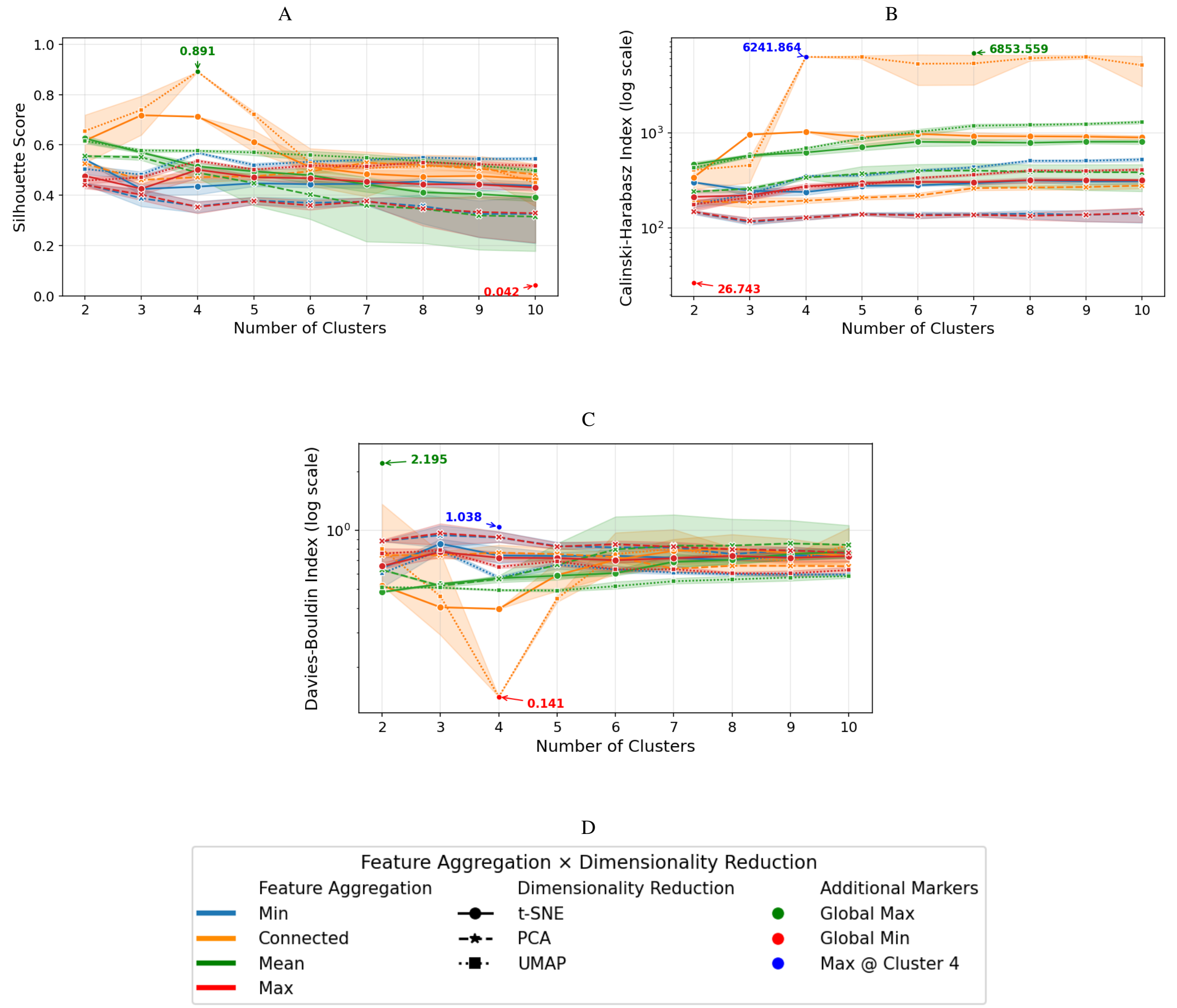


Clustering performance metrics across pipeline configurations and cluster counts. (A) Silhouette score vs. number of clusters ($k$), with global maxima (green) and minima (red) highlighted. (B) Calinski–Harabasz Index vs. $k$ (log scale). (C) Davies–Bouldin Index vs. $k$ (log scale). (D) Legend for metric curves. Across all metrics, the selected configuration (Connected Aggregation + UMAP + DEC at $k = 4$) provides the best balance of high silhouette, high CH, and low DB, indicated by the vertical dashed line. CH = Calinski–Harabasz Index; DB = Davies–Bouldin Index; DEC = deep embedded clustering; HCM = hypertrophic cardiomyopathy; UMAP = Uniform Manifold Approximation and Projection.

*Sensitivity Analysis: Phenogroup Stability After Exclusion of Patients with Prior SRT*

To address the concern that prior septal reduction therapy (SRT) may confound phenogroup assignments through post-intervention cardiac remodeling, we repeated the full *HCM-PhenotypeNet* pipeline after excluding the 12 patients with a documented history of SRT (all from Phenogroup A). The four-cluster partition was preserved in the SRT-excluded cohort ($n = 144$), with cluster quality metrics remaining high

**Supplemental Figure 2 Radar plot of normalized phenogroup features**

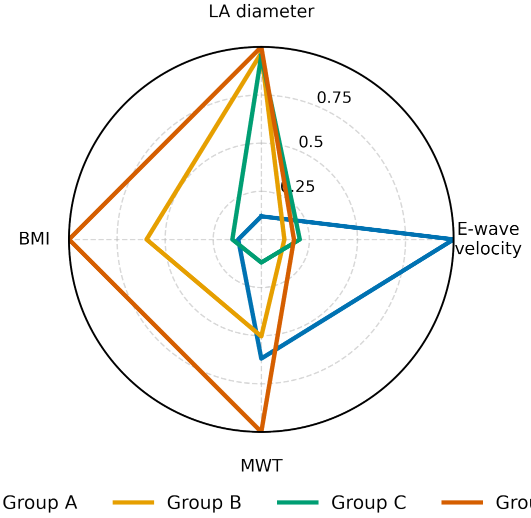


Radar plot showing normalized mean values (min–max 0.1–1.0) of four representative features—LA diameter, mitral E-wave velocity, MWT, and BMI—across the four HCM phenogroups (A–D) identified by the HCM-PhenotypeNet framework. Higher normalized values reflect greater structural, diastolic, or metabolic remodeling. Phenogroup A demonstrates marked LA enlargement and increased BMI; B shows intermediate remodeling with mild diastolic change; C exhibits mild hypertrophy with preserved filling; and D shows the greatest MWT and lowest BMI, representing a Hypertrophic Heart Failure-predominant phenotype. BMI = body mass index; HCM = hypertrophic cardiomyopathy; LA = left atrium; MWT = maximal wall thickness.

(silhouette ≈0.87; CH index ≈5,940; DB index ≈0.15). The composition and defining clinical characteristics of Phenogroups B, C, and D were unchanged. Within the reduced Phenogroup A ($n = 14$), the distinguishing features of larger left atrial diameter, higher BMI, and greater CAD prevalence remained intact. These findings confirm that the phenogroup structure is not an artifact of prior intervention history and that SRT status is a phenotypic consequence rather than a driver of cluster assignment.

*Association Rule Analysis*

This section expands on the main manuscript Section 4.5 (Interpretable Patterns via Association Rules). We applied association rule mining within each phenogroup (A–D) to characterize co-occurring clinical and echocardiographic imaging features. Continuous variables were binarized at prespecified clinical thresholds (e.g., BMI, IVS, age). To reduce false positives, missing clinical measurements were treated as not meeting threshold criteria (i.e., “absent” unless clinically documented). For association-rule mining, “Arrhythmia Events” was defined as the occurrence of any documented arrhythmic outcome on the Outcomes sheet, including atrial fibrillation, non-sustained VT, VT, cardioversion, ICD shock, or resuscitated cardiac arrest. Frequent itemsets were mined using FP-Growth, and rules were retained when *lift* $\geq 1.0$ [14]. Given our modest cohort and event counts, our use of associative rules is exploratory/hypothesis-generating.

*Association Rule Metrics*

For a rule $A \rightarrow B$ in a given phenogroup of size $N$:

- **Support** ($P(A \wedge B)$): fraction with both antecedent and consequent, $\text{Support} = \frac{n_{\text{both}}}{N}$.
- **Confidence** ($P(B|A)$): probability of the consequent given the antecedent, $\text{Confidence} = \frac{n_{\text{both}}}{n_{\text{ante}}}$.
- **Lift**: enrichment vs the phenogroup prevalence of the consequent, $\text{Lift} = \frac{\text{Confidence}}{\text{Consequent support}}$.

Counts were computed as: $n_{\text{both}} = \text{Support} \times N$, $n_{\text{ante}} = \text{Antecedent support} \times N$, $n_{\text{cons}} = \text{Consequent support} \times N$, where $N$ is the phenogroup size (A = 26, B = 59, C = 23, D = 48). Metrics were applied directly in Supplemental Table 5 (e.g., confidence = $n_{\text{both}}/n_{\text{ante}} = 11/17$ (64.7%).

*Association Rule Patterns by Phenogroup*

**Phenogroup A (Advanced obstructive/metabolic phenotype).** This phenogroup shows co-expression of obstructive physiology and cardiometabolic risk (highest SRT and mitral valve surgery rates).

- *{History of SRT}* $\rightarrow$ *{MR}* (confidence = 1.00; lift = 1.44; Supplemental Table 5, R1). **Finding:** All patients with prior SRT had MR. **Context:** In obstructive HCM, SAM-mediated MR is common, and comprehensive mitral valve assessment is recommended when SRT is considered [15]. **Cohort anchor:** Phenogroup A had the highest SRT (23%) and concomitant mitral valve surgery (8%), consistent with guideline thresholds for invasive therapy in symptomatic patients with significant gradients [15]. **Interpretation (hypothesis-generating):** The SRT$\rightarrow$MR link reflects treated obstructive physiology with residual/associated MR burden.

- *{Hypertension + BMI* $\geq 30$*}* $\rightarrow$ *{IVS* $\leq 13$ *mm (CMR)}* (confidence = 0.83; lift = 1.14; Table 5, R2). **Finding:** Obesity and hypertension clustered with only mild septal thickness. **Context:** In adults, HCM is typically diagnosed at maximal wall thickness $\geq 15$ mm (13–14 mm in relatives/with a pathogenic variant), situating IVS $\leq 13$ mm as mild [15]. **Interpretation (hypothesis-generating):** Cardiometabolic load may contribute to symptoms and remodeling without extreme hypertrophy, consistent with an obstructive/metabolic profile.

- *{Male + hyperlipidemia}* $\rightarrow$ *{history of CAD}* (confidence = 0.75; lift = 3.27; Table 5, R3). **Finding:** Dyslipidemia in males associated with greater CAD prevalence (27% vs 2% in Phenogroup D; Table 1). **Context:** Traditional cardiometabolic risks are common in HCM and warrant risk-factor modification; CAD evaluation is considered when results would alter management [15]. **Interpretation (hypothesis-generating):** A metabolic–atherosclerotic profile coexists with obstructive physiology in this phenogroup.

**Phenogroup B ("Intermediate, Non-obstructive HCM" phenotype):** This phenogroup is younger (mean age $\approx$ 51.6 y) and largely non-obstructive ( prior SRT uncommon [3%] $\approx$ 15% with any LVOT obstruction). Rule patterns indicate modest structural remodeling with atrial–valvular coupling.

- *{LA diameter* $\geq 40\,mm$*}* $\rightarrow$ *{MR}* (confidence = 0.81; Table 5, R4). **Finding:** In Cluster B (mean LA $\approx$ 38 mm; no prior valve interventions), LA $\geq$ 40 mm frequently co-occurred with MR. **Interpretation (hypothesis-generating):** LA enlargement likely reflects hemodynamic load from MR rather than diffuse myopathic progression.

- *{SAM}* $\rightarrow$ *{MR}* (confidence = 0.86; Table 5, R5). **Finding:** When SAM was present, MR co-occurred in most cases despite the group being largely non-obstructive and without prior septal intervention. **Context:** SAM is a recognized mechanism of MR in HCM [15]. **Interpretation (hypothesis-generating):** Even modest or provoked SAM may produce clinically relevant MR in early-stage disease.

- *{IVS* $\leq 13\,mm$ *+ LA diameter* $\geq 40\,mm$*}* $\rightarrow$ *{male}* (confidence = 0.87; Table 5, R6). **Finding:** Disproportionate LA enlargement with only mild septal thickening occurred predominantly in males. **Interpretation (hypothesis-generating):** Possible sex-related differences in atrial remodeling; cite supporting literature if expanded.

**Phenogroup C (Late-onset, mild non-obstructive HCM phenotype).** This phenogroup is older (mean age $\approx$ 59 y), shows modest structural remodeling, and has low LVOT obstruction prevalence (13%) and no prior SRT.

- *{Hypertension + MR (echo)}* $\rightarrow$ *{obesity (BMI* $\geq 30$*)}* (confidence = 0.86; support = 0.26; lift = 1.41; Table 5, R7). **Finding:** In Cluster C ($n$=23), 6/7 patients with {hypertension & MR} were obese. **Interpretation (hypothesis-generating):** Hypertension with MR concentrates in those with obesity, consistent with a metabolic–diastolic burden; emphasize weight and blood-pressure management.

- *{Female + hypertension}* $\rightarrow$ *{MR (echo)}* (confidence = 0.86; Table 5, R8). **Finding:** Women with hypertension frequently had MR ($\approx$ 71%); among hypertensive patients with MR, most were obese ($\approx$ 85%). **Interpretation (hypothesis-generating):** Pattern consistent with metabolic–diastolic load contributing to secondary MR and atrial enlargement in a mild structural phenotype.

- *{LA diameter* $\geq 40\,mm$ *+ age* $\geq 55\,y$*}* $\rightarrow$ *{first-degree family history of SCD}* (confidence = 0.63; lift $\approx$ 2.88; Table 5, R9). **Finding:** Co-occurrence was 5/8 (62.5%) among those meeting both criteria vs 5/23 (21.7%) cluster-wide. **Interpretation (hypothesis-generating):** Even in a mild, non-obstructive phenotype, older patients with atrial enlargement and a positive first-degree family history may merit heightened arrhythmia vigilance.

**Phenogroup D (Hypertrophic Heart Failure-predominant phenotype).** This phenogroup is middle-aged (mean age $\approx$ 53.0 y) and demonstrates a hypertrophic profile with marked septal hypertrophy (MWT $\approx$ 19.8 mm), a measurable propensity to obstruction ($\approx$ 19% with any LVOT obstruction; prior SRT occurred in 8%; no valve surgery), and scar-associated arrhythmic risk.

- *{SAM on echo}* $\rightarrow$ *{MR}* (12/14; confidence $\approx$ 0.86; Table 5, R10). **Finding:** When SAM was present, at least moderate MR co-occurred in most cases. **Context:** In obstructive HCM, SAM commonly produces MR via leaflet malcoaptation; MR contributes to symptom burden [15]. **Interpretation (hypothesis-generating):** The SAM$\rightarrow$MR pattern underscores dynamic LVOTO physiology in this phenogroup.

- *{LGE (CMR)}* $\rightarrow$ *{arrhythmic events}* (11/17; confidence $\approx$ 0.65; Table 5, R11). **Finding:** LGE-positive patients had more arrhythmic events. **Context:** Extensive LGE is associated with increased risk of life-threatening ventricular arrhythmias; in some studies, $\geq$ 15% LV mass is used as a high-risk threshold, and LGE can arbitrate ICD decisions when overall risk is uncertain [15]. **Interpretation (hypothesis-generating):** Scar burden may flag higher arrhythmic susceptibility in this phenogroup.

- *{LGE (CMR)} → {IVS ≥ 15 mm (CMR)}* (support = 0.21; confidence = 0.59; lift = 1.88; Table 5). **Finding:** Among LGE-positive patients, 10/17 (58.8%) had IVS ≥ 15 mm (cluster baseline 15/48, 31.3%). **Context:** In adults, maximal end-diastolic wall thickness ≥ 15 mm anywhere in the LV supports an HCM diagnosis; CMR enables accurate thickness/chamber measurement [15]. **Interpretation (hypothesis-generating):** Co-occurrence of hypertrophy and myocardial scar suggests a more advanced hypertrophic expression and supports guideline-directed SCD risk stratification.

Note: Counts are small and continuous variables were binarized; associations are exploratory.

Supplemental Table 5: Association Rules by Phenogroup Metrics.

| Rule ID | Phenogroup | Rule (Antecedent → Consequent)[a] | $n_{\text{both}}$ | $n_{\text{ante}}$ | $n_{\text{cons}}$ | Support (%) | Confidence (%) | Lift |
|---|---|---|---|---|---|---|---|---|
| R1 | A | {History of SRT} → {MR on Echo} | 6 | 6 | 18 | 23.08 | 100.00 | 1.44 |
| R2 | A | {Hypertension + BMI ≥30} → {IVS ≤13 (CMR)} | 10 | 12 | 19 | 38.46 | 83.33 | 1.14 |
| R3 | A | {Hyperlipidemia + Male} → {CAD history} | 6 | 8 | 7 | 23.08 | 75.00 | 3.27 |
| R4 | B | {LA Diameter ≥40 (Echo)} → {Mitral Regurg (Echo)} | 17 | 21 | 37 | 28.81 | 80.95 | 1.29 |
| R5 | B | {SAM on Echo} → {MR on Echo} | 12 | 14 | 37 | 20.34 | 85.71 | 1.36 |
| R6 | B | {IVS ≤13 (CMR) + LA ≥40 (Echo)} → {Male} | 13 | 15 | 32 | 22.03 | 86.67 | 1.62 |
| R7 | C | {Hypertension + MR on Echo} → {BMI ≥30} | 6 | 7 | 14 | 26.09 | 85.71 | 1.41 |
| R8 | C | {Female + Hypertension + BMI ≥30} → {MR on Echo} | 5 | 7 | 10 | 21.74 | 71.43 | 1.64 |
| R9 | C | {LA ≥40 (Echo) + Age ≥55} → {1st-degree SCD history} | 5 | 8 | 5 | 21.74 | 62.50 | 2.88 |
| R10 | D | {SAM on Echo} → {MR on Echo} | 12 | 14 | 27 | 25.00 | 85.71 | 1.52 |
| R11 | D | {LGE (CMR)} → {Arrhythmia Events} | 11 | 17 | 27 | 22.92 | 64.71 | 1.15 |
| R12 | D | {LGE (CMR)} → {IVS ≥15 (CMR)} | 10 | 17 | 15 | 20.83 | 58.82 | 1.88 |

[a] Association rules summarize frequent antecedent-to-consequent patterns within each phenogroup. Rules with antecedents and consequents indicate clinical patterns that co-occur within clusters. Support shows the proportion of patients meeting both antecedent and consequent; Confidence shows the proportion of antecedent-positive cases also meeting the consequent; Lift indicates the ratio to baseline prevalence. **Abbreviations:** BMI = body mass index; CAD = coronary artery disease; CMR = cardiac magnetic resonance; Echo = echocardiography; HCM = hypertrophic cardiomyopathy; IVS = interventricular septum; LA = left atrium; LGE = late gadolinium enhancement; MR = mitral regurgitation; SAM = systolic anterior motion; SCD = sudden cardiac death; SRT = septal reduction therapy.

*Future Directions, Clinical Integration, and Limitations*

This section contains the full text moved from the main manuscript Sections 4.6 and 4.7 to reduce main-text length.

*6.1 Future Directions and Clinical Integration*

This section expands the Future Directions and Clinical Integration content summarized in the main manuscript (Section 4.6).

Our study is a proof-of-concept that ML-driven multimodal phenotyping can identify meaningful HCM subgroups. However, further work is needed to translate this into practice. First, external validation on larger, multi-center cohorts is crucial to ensure these phenogroups generalize and to refine cluster definitions (our single-center results might not capture all HCM phenotypes). Federated learning or collaborations could build a more generalizable model without pooling raw data. Second, prospective studies should evaluate whether phenogroup membership adds prognostic value beyond traditional risk factors. For example, determining whether the "Advanced Obstructive/Metabolic" cluster predicts higher long-term AF or HF hospitalization rates could solidify the clinical utility of these phenogroups.

Integration into clinical workflows will also require addressing practical issues: model transparency, clinician acceptance, and ease of use. A system like *HCM-PhenotypeNet* could be embedded in echo lab software to automatically generate a phenogroup label when an HCM study is performed. This label—along with succinct explanations (e.g., "high-BMI, high-LA phenotype")—could alert the care team to particular risks or management pathways, and support the consideration of earlier interventions (such as referral for septal reduction therapy in a patient flagged as "Advanced Obstructive/Metabolic"). Such integration would require prospective validation before routine clinical adoption. Ensuring clinicians trust and understand the system will be key; the interpretability provided by association rules and clear phenogroup characteristics will help in this regard [16].

Finally, our work aligns with the push for precision medicine in cardiology. By demonstrating how dynamic imaging and clinical data can be combined to subclassify HCM, we pave the way for more personalized management strategies. With further refinement and validation, video-based phenotyping could guide tailored monitoring (e.g., more frequent Holter monitoring for Cluster A patients) and personalized therapy (e.g., early ICD consideration in certain phenogroups), ultimately improving outcomes for HCM patients.

Together, these steps outline a practical path from proof-of-concept phenogroup discovery to externally validated, clinically deployable decision support.

*6.2 Limitations*

This section provides a full discussion of the limitations that correspond to the concise statement included in the main manuscript (Section 4.6). This study has several limitations. It was retrospective and single-center, with a relatively small sample in some clusters (e.g., only 26 patients in Cluster A), which limits power to detect outcome differences and might not represent the full HCM spectrum. Only about half of patients underwent genetic testing, with uneven coverage across phenogroups; accordingly, genetic yields are descriptive and may reflect ascertainment (selection) bias. Our model was not tested on an independent dataset, so generalizability remains to be proven. Additionally, while unsupervised learning revealed clear clusters, the clinical interpretability of these data-driven groups—though enhanced by association rules—will require further expert validation. The association rule analysis itself has limitations: it binarized continuous data, possibly oversimplifying some variables, and it emphasizes frequent patterns (potentially missing rare but important associations). Diastolic function was summarized using a single mitral inflow parameter rather than a comprehensive diastology profile (e.g., E/e′), and more detailed diastolic phenotyping will be important in future work. We did not comprehensively characterize all imaging-based morphologic patterns of HCM, and future work could incorporate more detailed structural descriptors to refine these phenogroups. Finally, the phenogroup labels are descriptive rather than prescriptive; in particular, Phenogroup D is labeled the Hypertrophic Heart Failure-predominant phenotype to reflect its cardinal features—marked septal thickness, lean body habitus, and dominant HF symptom burden—rather than to imply a single genetic mechanism or canonical HCM subtype.

Moreover, interventions (like ICD implants or myectomy) may have reduced outcome differences between phenogroups. Lastly, implementing such a pipeline in real time would entail technical challenges and require prospective evaluation of its impact on clinical decision-making.

Despite these limitations, our findings illustrate the feasibility and utility of ML-based HCM phenotyping. Addressing the limitations—through larger studies, external validation, and prospective trials—will be an important next step to fully realize the potential of *HCM-PhenotypeNet*.

Future work will include multi-center external validation to assess the generalizability of the identified phenogroups across diverse patient populations.

Despite these limitations, the findings support the feasibility of automated multimodal phenotyping in HCM and motivate larger, multi-center validation and prospective evaluation. A further limitation specific to the deep learning component concerns model generalizability under distribution shift. The *HCM-Dynamic-Echo* feature extractor was fine-tuned on 1,553 video clips from a single academic center. Although pretraining on the large EchoNet-Dynamic corpus (Stanford University; >10,000 echocardiographic videos) provides a strong initialization, domain shift remains a concern: differences in ultrasound hardware, acquisition protocols, and post-processing pipelines across institutions can alter pixel-level spatiotemporal statistics in ways that may affect embedding quality without altering clinical diagnosis. Federated learning or cross-center harmonization strategies—such as instance normalization of video inputs or multi-site fine-tuning—should be explored in future work to ensure that phenogroup assignments generalize across imaging environments.
